# Mutations in *ZBTB20* in individuals with persistent stuttering

**DOI:** 10.1101/2022.11.03.22281471

**Authors:** Carlos E Frigerio Domingues, Muhammad Hashim Raza, Tae-Un Han, Terra Barnes, Philip Shaw, Gustavo Sudre, Sheikh Riazuddin, Robert J. Morell, Dennis Drayna

## Abstract

**Background:** Previous studies identified a strong linkage signal for non-syndromic persistent developmental stuttering on chromosome 3q13.2-3q13.33 in a large consanguineous family. To identify the causative genetic variant at this locus, including we performed further analysis, including whole exome, whole genome, and targeting Sanger sequencing.

**Results:** We identified a homozygous rare c.2155G>A variant in *ZBTB20* in individuals who stutter. This mutation encodes Isoleucine in place of a highly conserved Valine at amino acid position 719 in ZBTB20 that co-segregates (LOD = 4.23) with stuttering under a model of recessive inheritance with reduced penetrance in this family. Coding variants in this gene were significantly more frequent in a multiethnic cohort of unrelated individuals who stutter than in individuals in large population databases comprised of our normal control subjects and gnomAD database subjects matched for ethnicity. *ZBTB20* encodes a zinc-finger transcription factor, and luciferase reporter constructs using genes known to be regulated by ZBTB20 showed that the Ile719 mutant form of the protein displays altered transcriptional regulation *in vitro*. Although homozygosity for mutations in *ZBTB20* has not previously been observed, dominant mutations in *ZBTB20* have been reported in Primrose syndrome, a rare Mendelian dominant disorder characterized by tall stature, developmental delay, dysgenesis of the corpus callosum, but generally not speech disorders. Clinical re-examination of the affected family members ten years after their original ascertainment revealed only some suggestive signs associated with Primrose syndrome.

**Conclusions:** Our findings support variants in *ZBTB20* as a cause of stuttering in a large family, and rarely in the wider population. Previous studies in mice have demonstrated that *Zbtb20* is required for the development of astrocytes, a brain cell type previously implicated in an animal model of stuttering. Our findings support our hypothesis that astrocyte pathology is involved in this disorder. Our findings also broaden the medical genetic view of disorders of *ZBTB20*, and demonstrate that individuals carrying a homozygous mutation in this gene can be viable, and that they can primarily display persistent developmental stuttering.

## Introduction

Stuttering is a common neurodevelopmental speech disorder that affects the fluency of speech, and is characterized by repetitions and prolongations of speech sounds, and by silent interruptions in the flow of speech known as blocks [1,2]. Twin and adoption studies indicate a high heritability for this disorder [3], however Mendelian segregation does not typically occur [4] and stuttering is considered to be a complex genetic disorder. Linkage studies in consanguineous and polygamous families have identified a number of loci that display strong linkage to this disorder [5,6,7,8, 9], and causative genes for stuttering have been identified at a number of these loci [10,11]. The products of the genes identified thus far all physically interact and participate in functions important to intracellular trafficking. Animal studies aimed at elucidating the cellular and anatomic neuropathology caused by these mutations have recently revealed deficits in vocalization and abnormalities in astrocytes, particularly in the corpus callosum [12,13].

We have previously reported a large consanguineous family in which many members are affected by persistent neurodevelopmental stuttering. Linkage analysis identified significant linkage on chromosome 3 under an autosomal recessive model with reduced penetrance [8]. Here we have performed additional linkage analyses as well as whole exome, whole genome, and Sanger sequencing, which revealed a unique rare homozygous coding variant in *ZBTB20* (MIM: 606025) that co-segregates with stuttering in this family. Although no individuals with homozygous mutations in *ZBTB20* have previously been observed, heterozygous mutations have been associated with Primrose syndrome, a rare multisystem developmental disorder [14, 15, 16, 17, 18, 19, 20, 21, 22, 23].

*ZBTB20*, also known as *DPZF* [24], *HOF* [25], or *ZNF288* is a member of the POK (POZ and Krüppel) family of transcription factors. We sought to determine if this variant exerted a functional effect on transcription using a luciferase reporter construct in transfected cells. We also performed additional clinical evaluations of the affected subjects in this family, and constructed and tested ultrasonic vocalizations in a knock-in mouse line that carries this mutation.

## Results

### Stuttering in Family PKST77

Consanguineous family PKST77 has been previous described [8]. Speech abnormalities in all five affected subjects were consistent with typical, non-syndromic persistent developmental stuttering. Based on our subsequent genetic findings, family members were re-evaluated 10 years after their initial enrollment. Clinical examinations of four available affected family members were performed at the Allama Iqbal Medical College, Lahore, Pakistan, and included assessment of medical history, physical examination, neurological evaluation. Brain MRI scans performed in Pakistan were analyzed in the Neurobehavioral Clinical Research Section, National Human Genome Research Institute.

### Identification of variants in *ZBTB20*

The pedigree of Family PKST77 is shown in Figure 1A. This family displays highly significant genetic linkage between persistent stuttering and markers on chromosome 3q [8]. We re-investigated the previously reported linkage using a genome-wide SNP linkage scan under a model of recessive inheritance with reduced penetrance [8], which confirmed significant linkage scores in this region and nowhere else across the genome. As previously observed with lower density markers, assignment of precise linkage boundaries was limited by instances of apparent non-Mendelian segregation, consistent with the accepted nature of stuttering as a complex trait. Whole-genome sequencing was performed in 16 individuals (III-13, III-17, III-21, III-22, IV-23, IV-24, IV-25, IV-26, IV-27, IV-28, IV-29, IV-30, IV-31, IV-32, IV-33, IV-34) within this family. The whole-genome sequencing quality control followed the minimum standards requirements established by The American Genome Center at Uniformed Service University, with at least a minimum 30X Illumina coverage, 75% Q30 scores, and 100 gigabases of sequence produced per sample. The analysis identified a mean of 3,824,970 SNVs (range 3,745,519 – 3,932,504 variants) and a mean of 1,023,407.56 indels (range 998,694 – 1,051,753 indels) (Suppl.1.1).

**Figure 1.**
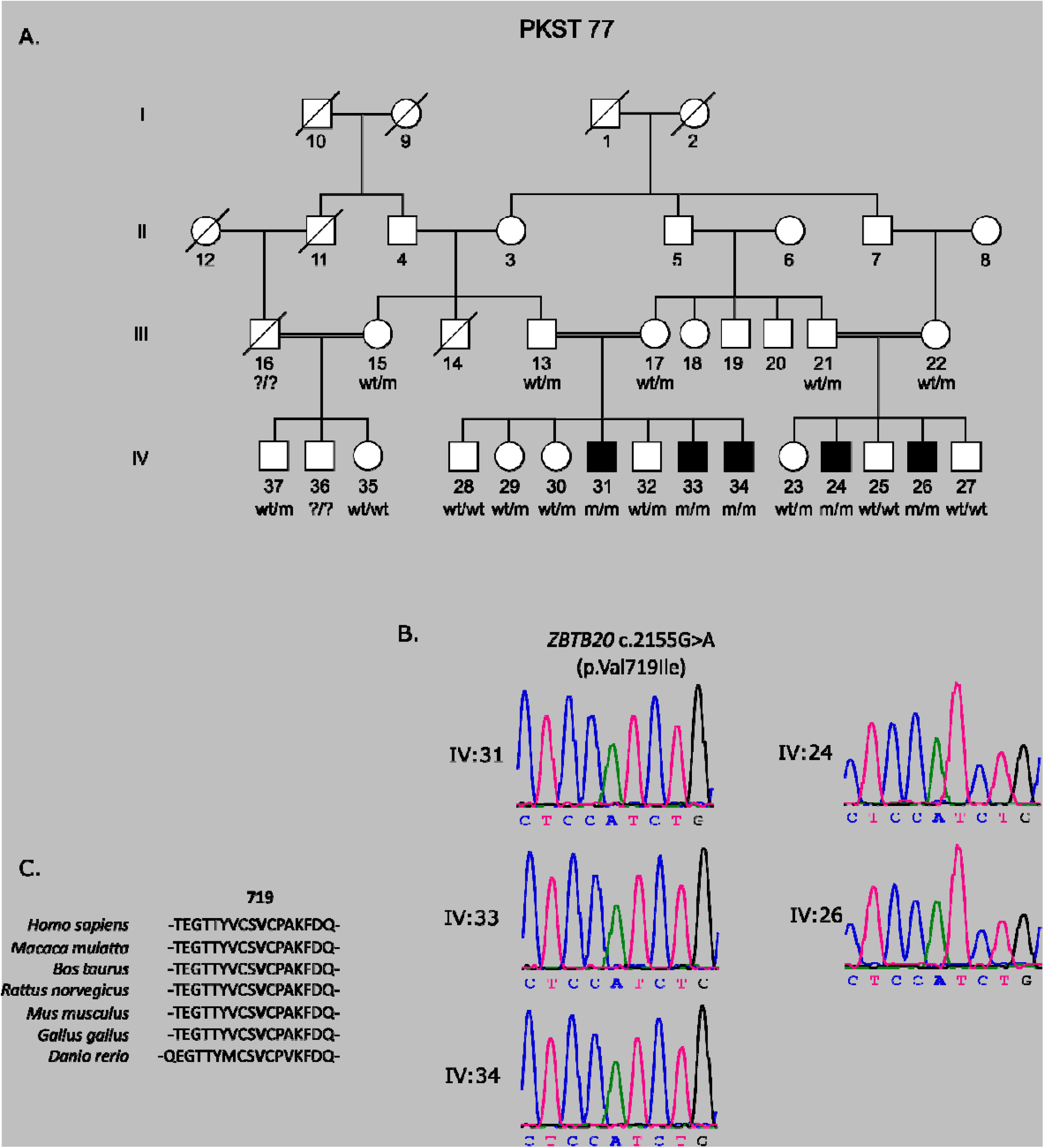
**A**. Pedigree of stuttering family PKST77. Individuals diagnosed as affected represented by filled symbols. wt, wildtype allele; m, mutated allele; wt/wt, homozygote wildtype; wt/m, heterozygote mutated; m/m, homozygote mutated B. Dideoxy Sanger sequencing traces showing the c.2155G>A variant in ZBTB20 in available family members. C. Amino acid alignments across species of the Valine 719, in bold

Once annotated, all variants underwent a bioinformatics sorting analysis as follows. First, variants not on chromosome 3 were eliminated, leaving 12,283 variants (SNPs = 10,181; Indels = 2,102). Next, variants residing outside the region of significant linkage (11,925 variants) were excluded, leaving a total of 358 variants, all residing withing the previously demonstrated linkage region. Many of these occurred commonly (allele frequency ≥ 0.01) in gnomAD v2.1 (Genome Aggregation Database), the 1000 Genomes phase 3 dataset, or in the NHLBI ESP6400 dataset. These were eliminated from consideration, and variants with minor allele frequency (MAF) < 0.01 in at least one of these databases were considered further. Of the remaining 41 variants, we focused on the 17 coding variants (15 nonsynonymous, 1 frameshift deletion, and 1 stop gain, Table 1), based on the hypothesis that the causative allele in PKST77 could be a coding allele of large effect. Only one of these 17 rare variants co-segregated with the disorder under the expected recessive model observed within the family. The variant (c.2155G>A) was in *ZBTB20* (GenBank: NM_001164342). It occurs at the beginning of exon 5 and is predicted to cause a valine-to-isoleucine substitution (p.Val719Ile), and the location is illustrated in Figure 2. The wild-type valine is conserved across all vertebrates with the exception of coelacanth (*Latimeria chalumnae)*, which carries a phenylamine at this position (Figure 1C and Suppl.1.2). The Val719Ile change is predicted to be disease causing (score:29) by Mutation Taster (http://www.mutationtaster.org/); probably damaging (score: 0.999) by PolyPhen-2 (http://genetics.bwh.harvard.edu/pph2/); and tolerated (score: 0.14) by SIFT (https://sift.bii.a-star.edu.sg/).

**Table 1.**
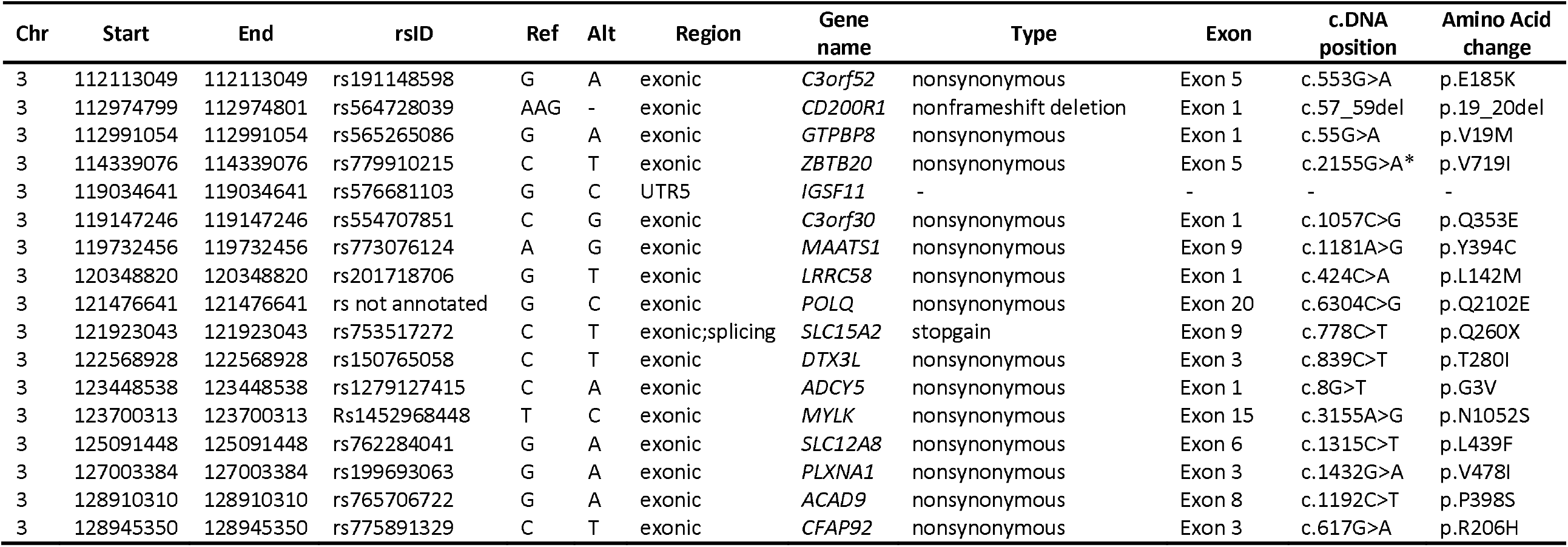
List of 17 coding variants (MAF < 0.01) under final investigation. **Note:** ^*^*ZBTB20* variant present in all affected individuals in homozygous state

**Figure 2.**
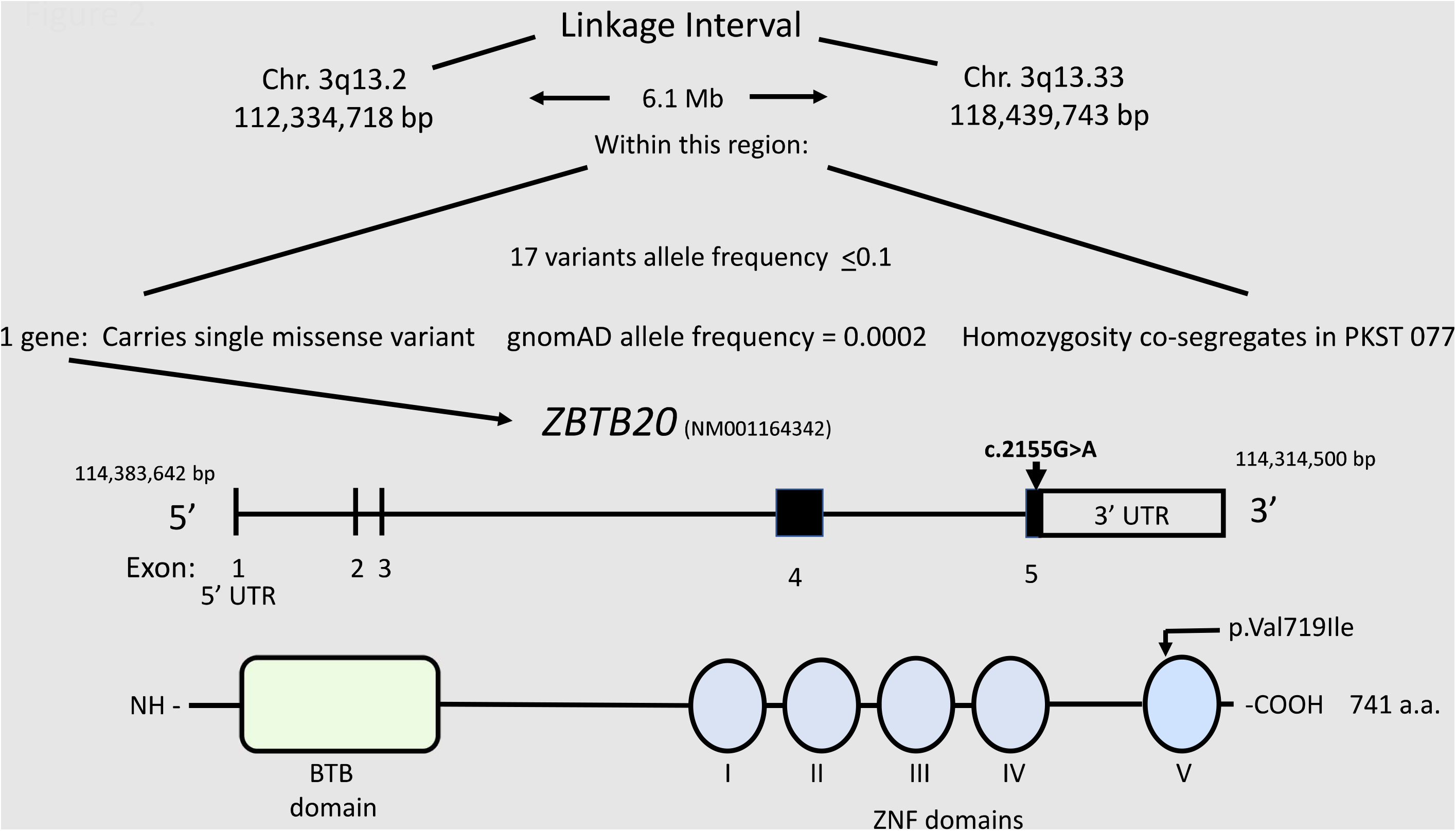
Chromosome 3 linkage region, structure of ZBTB20 gene, and mutation location

In all family members previously analyzed with whole genome sequencing, the presence of the homozygous variant c.2155G>A in *ZBTB20* was confirmed by Sanger sequencing as shown in Figure 1B. In addition, we performed dideoxy sequencing in five additional unaffected members. Across the individuals with fully confirmed genotype data, homozygosity for the Val719Ile variant co-segregates with persistent stuttering.

This mutation is rare. It is present in 22/141,456 individuals in the gnomAD Database (v.2.1.1), and all 22 of these individuals are heterozygous. We note that the individuals in the gnomAD database are of unknown speech phenotype (https://gnomad.broadinstitute.org/variant/rs779910215?dataset=gnomad_r2_1). A total of 13 other missense variants were found in gnomAD, all present as heterozygotes. This indicates that the affected individuals in PKST77 are the only known individuals homozygous for a coding mutation in *ZBTB20*. We also analyzed our custom-built Communication Exome Database (CED) (N = 1,941 unrelated individuals), sorting for coding variants among stuttering individuals in different cohort populations (N = 1,074 individuals) and matched controls (N = 867 individuals). No occurrences of the *ZBTB20* c.2155G>A variant were identified.

To estimate the contribution of mutations in *ZBTB20* to stuttering overall, we performed a burden test that compared the rate of rare variants in affected and control individuals. Although the frequency of *ZBTB20* coding variants in our combined case groups (66/1,074) was nominally higher than that in the combined controls (41/867), the difference in frequency between cases and controls did not reach statistical significance in any of our population-specific case/control subgroups (Suppl.1.3). We also compared the frequency of rare coding variants in our stuttering cohort (2,148 chromosomes) with that in the general population represented in the gnomAD v.2.1 dataset, (282,912 chromosomes), which likely contains both unaffected and affected individuals (https://gnomad.broadinstitute.org/faq#what-populations-are-represented-in-the-gnomad-data). Here, the frequency of rare *ZBTB20* variants in our stuttering group was higher overall. We observed more variants in total in our combined stuttering group, which is highly ethnically diverse (88 alleles/2,148 total alleles) than in an ethnically diverse gnomAD population containing similar proportions of European, Asian, and African individuals (5,450 alleles/282,912 total alleles, Chi-square = 52,716, p = 3.85 × 10^−13^). These data indicate that rare coding variants of all types in *ZBTB20* are elevated in the stuttering populations.

### The ZBTB20_Val719Ile mutation suppresses transcriptional repressor activity of ZBTB20 in a human cell

ZBTB20 has been reported to act as a transcriptional repressor for multiple target genes including alpha fetoprotein (AFP) and inhibitor of kB alpha (IκBα) [26]. To investigate the functional effects of the ZBTB20_Val719Ile mutation on protein activity in human cells, we performed a reporter gene assay using the co-expression of ZBTB20 and luciferase proteins transcribed under control of the AFP promoter. The wildtype and Val719Ile variants of ZBTB20 were transiently over-expressed separately in the human HepG2 cells, along with AFP-luciferase vector, where the measured luciferase activity directly reflected the intracellular activity of ZBTB as a transcriptional repressor. The luciferase activity was reduced about 40% in both the low (20 ng) and high (40 ng) dosage of wildtype ZBTB20 transfection comparing transfection to a control vector. Transfection of the same amount of ZBTB20_Ile719 expression vector rendered the transcriptional repressor activity significantly suppressed compared to transfection of wildtype ZBTB20 in both low (*p* = 0.034) and high dosage (*p* = 0.0083). The data shown in Figure 3 demonstrates that the ZBTB20_Ile719 is associated with a decrease in the activity of the mutant ZBTB20 protein as a transcriptional repressor in human cells.

**Figure 3.**
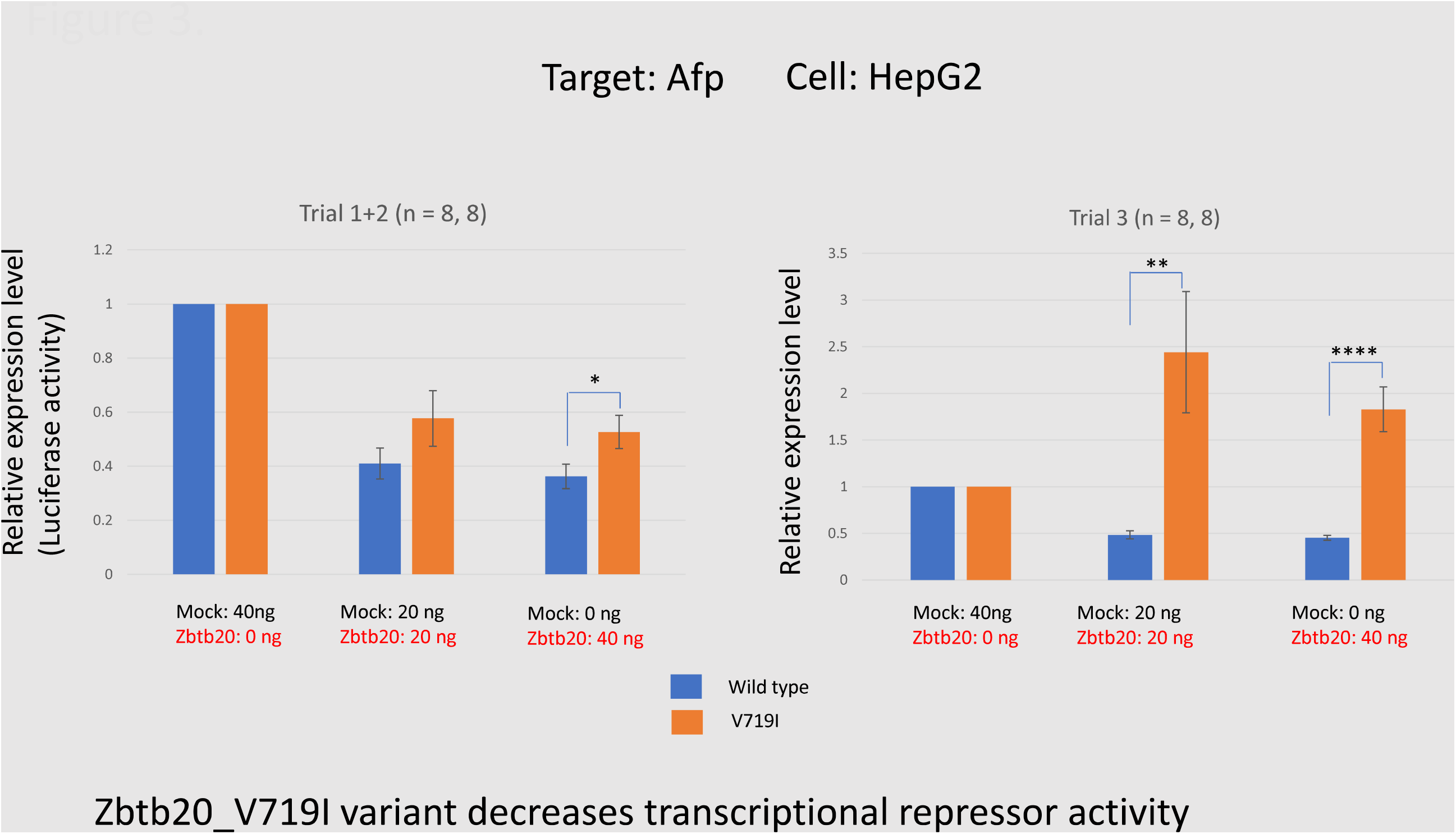
The ZBTB20_Val719Ile variant decreases transcriptional repressor activity. Human HepG2 cells were co-transfected with the pCMV6-ZBTB20 wild type or Val719Ile mutation, and the pGL4.10-AFP reporter vector expressing a luciferase gene under control of the AFP promoter (-2320 to +23). A Renilla-luciferase vector (pRL-TK) was also co-transfected to control for transfection efficiency. Relative expression level was calculated dividing luciferase activity of pCMV6-ZBTB20 by that of pCMV6_Empty vector as a mock control. A total of 16 replications in three independent experiments are shown (total n= 16, 16). Legend: Wt, wildtype ZBTB20; Error bar shows SEM.*p value = 0.034; **p value = 0.0083

### Ultrasonic mouse vocalizations

Animal models of stuttering have been created by generating mouse lines carrying a range of mutations in the *GNPTAB* gene found in humans who stutter [12, 13]. Such animals displayed altered ultrasonic vocalizations with abnormalities similar in some aspects to those observed in humans who stutter. We used CRISPR-cas to engineer a mouse line carrying the Val719Ile mutation in *Zbtb20* [27, Suppl.2.4]. Mice carrying the Zbtb20 Val719Ile variant displayed normal pup isolation call vocalization with respect to amount of vocalization and the spectral frequencies observed. The length of the silent intervals in both the inter-but and intra-bout vocalizations showed a suggestion of increased length compared to those in their genetically wild-type littermates, but this difference was not statistically significant (data not shown).

### Clinical evaluations

*De novo* heterozygous missense variants in *ZBTB20* have previously been associated with Primrose syndrome, a rare autosomal dominant disorder that is characterized by a slowly progressive condition associated with tall stature, increased head circumference, atypical facial features, cognitive deficits associated with autism spectrum disorder, and ectopic calcifications [28]. Primrose syndrome patients may subsequently begin to manifest other clinical features including distal muscle atrophy, hearing loss, cataracts, sparse body hair, disturbed glucose metabolism, dense calcification of external ears, and anatomic abnormalities of the corpus callosum. Although Primrose syndrome is clinically consolidated and recognizable in adults, the diagnosis in infants and children is considered difficult [29]. Four affected members of family PKST77 were available ten years after their original enrollment, and they were examined in greater detail (Tables 2 and 3, Suppl.1.4). Many of the most common features of Primrose syndrome, including tall stature and increased head circumference, were absent in these family members. However mild facial dysmorphism and, in some members, mild intellectual disability was noted.

**Table 2.**
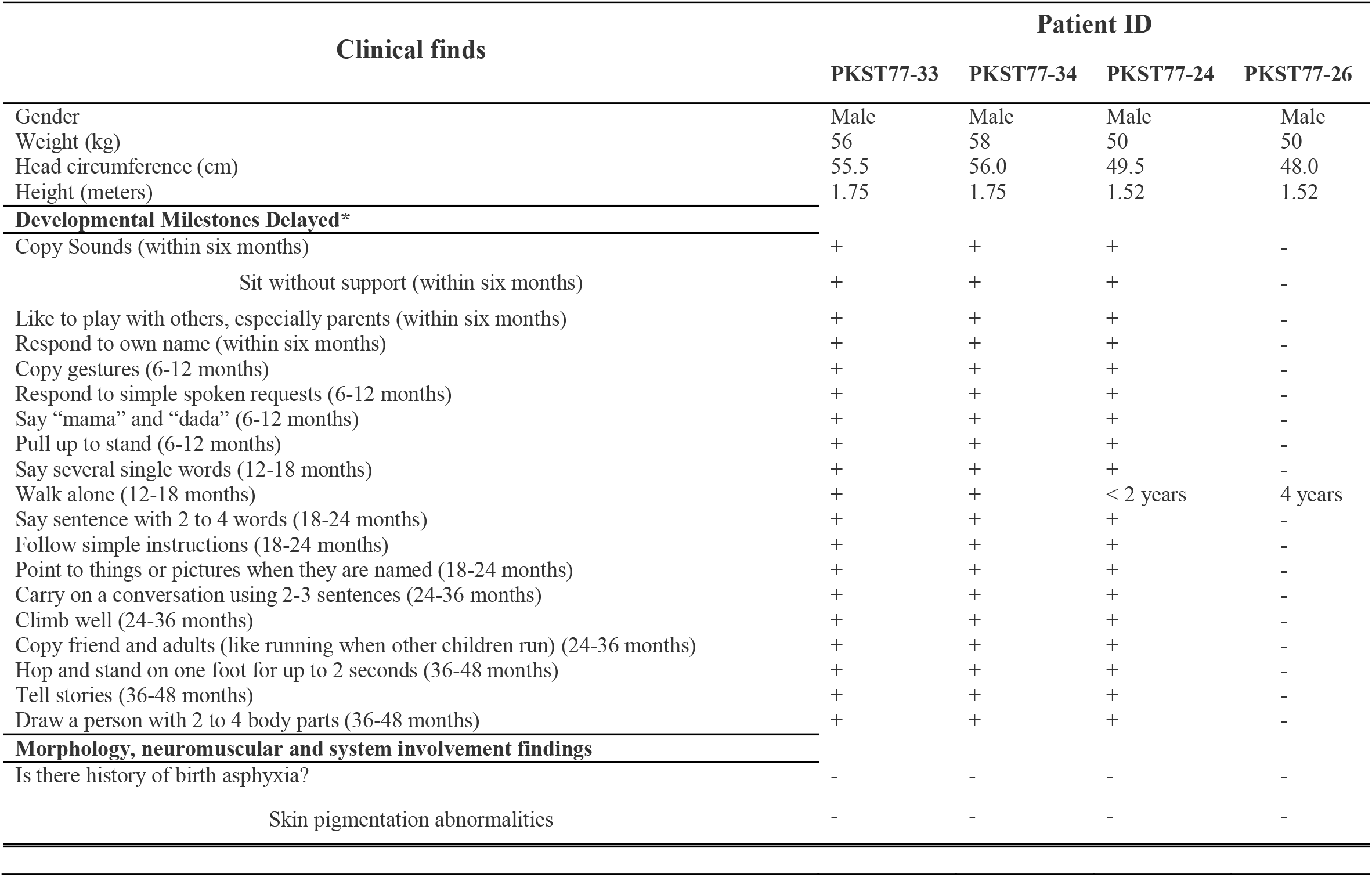
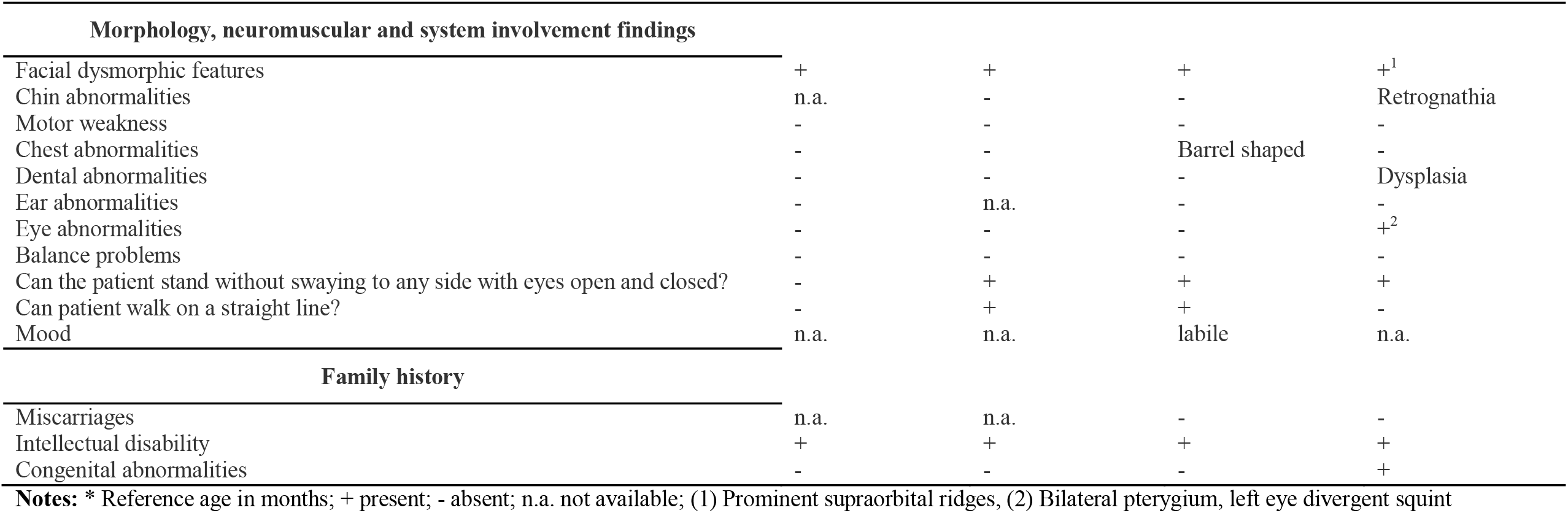
PKST 077 Clinical evaluation report.

**Table 3.** Structural characteristics of the Corpus Callosum in members of Pakistani stuttering family PKST077.

| **PKST77 Family**<br>**members** | **Corpus Callosum**<br>**Volume (mm<sup>3</sup>)** | **Corpus Callosum**<br>**Fraction Anisotropy**<br>**(FA, n.u.)** |
| --- | --- | --- |
| PKST77-24 | 3652.1 | 0.36 |
| PKST77-26 | 4994.3 | 0.32 |
| PKST77-27 | 3982.8 | 0.34 |

Brain MRI scans were available for three family members, one homozygous normal unaffected individual and two homozygous affected individuals. T1-weighted images (176 axial slices, 0.9-mm slice thickness, 256□×□256 acquisition matrix) and diffusion weighted images (TE=89ms, TR=11.1s, 128x128 matrix, slice thickness = 2 mm, gap = 2 mm, 102 directions) were acquired in a 3-T Siemens MAGNETOM Vida scanner. Freesurfer version 7.1.1 (https://surfer.nmr.mgh.harvard.edu) and FSL version 6.0.1 (https://fsl.fmrib.ox.ac.uk/) were used to process T1w and DWI data, respectively. Structural characteristics are shown in Table 2. While this sample size was too small to draw statistically significant conclusions, no striking differences were observed in the structure of the corpus callosum in these individuals.

In conclusion, the clinical phenotype information available from the members of PKST77 was limited in our follow-up evaluations. In these evaluations, we noted some individuals with mild facial dysmorphism and intellectual disability, although these were not consistent across affected individuals. Both of these signs are observed in Primrose Syndrome, but our limited data precludes a firm conclusion that this represents a variant form of this disorder.

## Discussion

Persistent stuttering has been shown to be highly genetic in origin, and several individual genes that contribute to this disorder have been described [10,11]. However, the genes found thus far appear to account for only a fraction of cases of stuttering, and it is clear that additional genetic factors exist. Data supporting the view that variants in *ZBTB20* cause of stuttering comes from four lines of evidence. First, strong evidence for linkage of stuttering to this region has been documented in a large, consanguineous family [8]. Second, homozygous mutations at a highly conserved site in the protein encoded by this gene co-segregate with stuttering under an appropriate model of recessive transmission with reduced penetrance, and no other variants in the linkage region passed our filtering criteria. Third, in a population-based burden test we found a higher frequency of rare variants in this gene in stuttering cases compared to ethnically matched control database groups. Fourth, this gene encodes a transcription factor. The variant in this gene present in the large consanguineous family causes a functional defect in its transcriptional regulatory activity in an *in vitro* luciferase reporter system. In addition to these four lines of evidence, *ZBTB20* is known to be a major determinant of astrocyte development in the brain [30, 31]. Studies have identified prominent astrocyte deficits in mice that exhibit stuttering-like vocalization deficits due to a mutation in *GNPTAB*, a previously identified stuttering gene [13]. Thus, our findings in *ZBTB20* may further support a role for astrocyte deficits in humans who stutter.

Primrose syndrome is a dominant disorder, and all patients described to date carry a mutation in just one of their two *ZBTB20* alleles. It is not yet known if these mutations act as dominant negatives or via haploinsufficiency. Given the involvement of ZBTB20 in a range of important developmental processes [30, 31, 32, 33, 34], it is possible that homozygosity for mutations in this gene is generally incompatible with viability. This idea is supported by the lack of individuals homozygous for any coding sequence mutation in *ZBTB20* in the existing gnomAD v.2.1 dataset, representing 141,456 multi-ethnic individuals.

We note that, in contrast to the homozygosity for the variant in family PKST77, our population-based burden test identified mostly heterozygous *ZBTB20* variants. This was the case for both our stuttering groups and control groups. We suggest that while the unusual homozygosity in family PKST77 resulted in identification of a genotype with large effect, other *ZBTB20* alleles may exert an effect in a heterozygous state, but discernable only at the population level. It is also noteworthy that stuttering has not been described in Primrose Syndrome to date (https://www.omim.org/entry/259050). We note that the stuttering genes identified thus far had been previously associated with other rare Mendelian medical genetic disorders, none of which had stuttering as a noted feature of their clinical presentation [10, 11, https://www.ninds.nih.gov/Disorders/Patient-Caregiver-Education/Fact-Sheets/Mucolipidoses-Fact-Sheet, https://omim.org/entry/607244]. The reasons for this are not fully understood, but for one of these genes, it has been shown that the mutational spectrum present in stuttering does not overlap with the mutations in this gene seen in the rare medical genetic disorder [35].

Zbtb20 is fundamental in brain development and function, and it is expressed in developing hippocampal neurons, as well as in astrocytes in the cerebral cortex and cerebellum [32, 33, 34] where it has been demonstrated to be an essential regulator of astrocyte development. During this development, Zbtb20 is highly expressed in neural precursor cells at late stages of neocortical development, and it is involved in the differentiation and maturation of astrocytes while suppressing the development of neurons [30].

Animal models of stuttering have been created by inserting human stuttering mutations into the mouse [12, 13] and such mice display abnormalities in their ultrasonic vocalizations that share similarities with the speech deficits in human who stutter and carry these same mutations. The mice display anomalies in their brain astrocytes, and this pathology is significantly displayed in the corpus callosum [13]. Structural abnormalities of the corpus callosum are a feature of Primrose syndrome [14, 20, 36], and although we did not observe such abnormalities in the members of family PKST077, the small number of subjects available for evaluation limits this conclusion. It is interesting to note that this gene plays a major role in the development of mammalian astrocytes [30], a cell type previously discovered in a mouse model of stuttering caused by another gene [13]. While our findings do not speak to the state of potential astrocyte pathology in the members of family PKST77, they suggest a functional deficit in the transcriptional regulatory function of ZBTB20 in this family. If astrocyte pathology is associated with human stuttering, our findings here would be consistent with this, and should warrant additional investigation of this so-called astrocyte hypothesis in human stuttering. We note these findings implicate astrocyte pathology indirectly, and that previous findings were found with mutations in a different gene (*GNPTAB*) using a mouse model of stuttering. The present findings in human stuttering, originally based on a mechanistically agnostic linkage study, demonstrate an alteration in function in a transcription factor necessary for astrocyte development.

We suggest that mutations in *ZBTB20* are a relatively rare cause of stuttering, as such mutations were observed in only a small number of unrelated individuals who stutter beyond family PKST077. Nevertheless, we believe these findings open new avenues of research into the neuropathological underpinnings of this enigmatic disorder.

## Materials and Methods

### Subjects

All human subjects, including individuals from a consanguineous family (PKST77), with the inclusion diagnosis of non-syndromic persistent stuttering were enrolled with written informed consent under NIH protocol 97-DC-0057 (Genetic studies of stuttering), also reviewed and approved by the IRB of the Centre of Excellence in Molecular Biology, University of the Punjab, as previously described (Raza et al., 2010). All subjects were age 8 and older.

Documented neurologically normal control DNA samples (NDPT; N = 460) were obtained from the National Institute of Neurological Disorders and Stroke (NINDS) panels NDPT006, NDPT020, NDPT023, NDPT079, NDPT082, NDPT093 and NDPT094 from the Coriell Cell Repository (available at https://www.coriell.org/Search?grid=0&q=ninds&csId=&f_0=NINDS+Repository&f_1=Control&f_2=96+well+plate+of+DNA+samples). Our study also included unrelated affected individuals (total N = 1,074) and population-matched control individuals (N = 867), which was made up of Pakistani affected individuals (PKSTR; N = 72) and Pakistani normally fluent controls (PKNR; N = 96), Cameroonian affected (STCR; N = 105) and control (RC; N = 93) individuals, North American affected individuals, including those from our NIH group (NIH; N = 739) and Brazilian affected individuals (BRMII; N = 158) and controls (BRCO; N = 218). Stuttering was diagnosed as previously described [37, 38]. Individuals were classified as affected as previously described [10]. Affected individuals were classified as unrelated by self-report, and no genotypic evidence for the relatedness among these subjects was observed. The identity of all subjects is protected under the privacy and confidentiality guidelines of the National Institutes of Health Institutional Review Board (Protocol 97-DC-0057), and the identities of the subjects are not known outside of the research group.

### Linkage analysis

Two independent linkage analyses were performed in family PKST77. The first was described by Raza et al. (2010). The second, newly performed in this study, used the same subjects, here genotyped at 551,839 SNPs assayed on the Illumina Infinium CoreExome-24 v.1.1 arrays according to manufacturer’s protocols. The linkage analysis was performed using MERLIN 1.1.2. (https://csg.sph.umich.edu/abecasis/Merlin/download/) with parametric and non-parametric models.

### Whole Exome and Genomic Sequencing

Genomic DNA was purified from whole-blood samples using a PAXgene^®^ Blood DNA Kit (QIAGEN), or from saliva samples using an Oragene^®^ DNA kit (DNA Genotek Inc., Canada), according to the manufacturer’s protocols. Exome enriched DNA libraries were prepared using Nextera Rapid Capture Exome (Illumina, San Diego, CA, USA) and were sequenced on an Illumina HiSeq1500 with 75 × 75 paired-end reads in 1,941 samples. Additionally, whole-genome libraries were prepared on 16 members of the family PKST77 using an Illumina Nextera TruSeq DNA PCR-free kit and sequenced on an Illumina HiSeq X Ten at The American Genome Center at Uniformed Service University (TAGC-USU, https://www.usuhs.edu/chirp/bioinformatics).

### Sequence Data Analyses

Whole-exome and whole-genome sequence quality control reads were aligned to Genome Reference Consortium Human Build 38 patch release 12 (GRCh38.p12).

### Bioinformatic pipelines

Whole-exome data was analyzed using the bcbio-nextgen germline variant pipeline (https://bcbio-nextgen.readthedocs.io/en/latest/). For variant calling, the “ensemble” vcf file was used, which required that a variant be detected in at least two of the following variant callers: gatk-haplotype, platypus, vascan, freebayes, or Samtools. The complete snakemake pipeline script is provided in the supplement (Suppl.2.1 and Suppl.2.2).

Whole-genome data: for genomic alignment was used NovoAlign - Novocraft 3.08.02 (http://www.novocraft.com/products/novoalign/), followed by Picard Toolkit 2.9.2 (https://broadinstitute.github.io/picard/), the variants were called using Genome Analysis Tool Kit 3.8.0 (GATK) (https://software.broadinstitute.org/gatk/) according to Best Practices Workflow (https://software.broadinstitute.org/gatk/best-practices/) and, called variants were annotated using ANNOVAR, release 2017Jul16. (http://annovar.openbioinformatics.org/en/latest/).

### Database annotation and variant sort

A total of 1,941 samples among unrelated stuttering cases and controls were analyzed in a custom-built Communication Exome Database (CED). This MySQL relational database included metadata on each individual, variants from the corresponding “ensembl.vcf” file, and extensive metadata on each variant including annotations from the Genome Aggregation Database (gnomAD v2.1) (https://gnomad.broadinstitute.org/), 1000 Genomes Project phase 3 (http://useast.ensembl.org/Homo_sapiens/Info/Index), National Heart, Lung, and Blood Institute – Exome Sequencing Project (NHLBI-ESP) Exome Variant Server (https://evs.gs.washington.edu/EVS/), Kaviar Project (http://db.systemsbiology.net/kaviar/cgi-pub/Kaviar.pl), dbSNP (https://www.ncbi.nlm.nih.gov/snp/). Variants with minor allele frequency _≥_ 0.01 were filtered out. The remaining variants, sorted according to chromosome position, gene function, type of amino acid changed, evolutionary conservation level and clinical information using annotations provided by public databases (including RefSeq [https://www.ncbi.nlm.nih.gov/refseq/], knownGene [http://genome.ucsc.edu/] dbNSFP database, [https://sites.google.com/site/jpopgen/dbNSFP] and ClinVar database [https://www.ncbi.nlm.nih.gov/clinvar/]).

### Sanger sequencing

Sanger sequencing was performed to validate all variants previously identified by whole exome sequencing. Oligos were designed using Primer3 (http://bioinfo.ut.ee/primer3-0.4.0/) and are listed in the supplemental materials (Suppl.2.3). Sanger dideoxy sequencing of all exons and their 40 bp flanking regions was performed using BigDye^®^ Terminator Cycle Sequencing Kit v3.1 (Applied Biosystems, Foster City, CA, USA) with standard manufacturer’s protocols. Sequence reactions were resolved on an ABI Genetic Analyzer 3730xl instrument (Applied Biosystems) and the electropherograms were evaluated using DNASTAR^©^ Lasergene SeqMan Pro™ (v.13.0.2) (https://www.dnastar.com/software/lasergene/).

### Construction of *ZBTB20* expression vectors

*ZBTB20* expression vector was prepared using OriGene (OriGene Tech. Inc., MD), TrueORF^®^ cDNA clone (Cat#: RC228567) (available in: https://www.origene.com/catalog/cdna-clones/expression-plasmids/rc228567/zbtb20-nm_001164342-human-tagged-orf-clone). This vector expresses *ZBTB20* human isoform 1 (NM_001164342) under the control of a CMV promoter. The wild type valine at position 719 of ZBTB20 was replaced with isoleucine by introducing a point mutation using QuickChange II Site-Directed Mutagenesis kit (Agilent Tech. Inc., CA). The promoter of alpha fetoprotein (*AFP*, -2320 to 23 bp) was amplified from genomic DNA of a normal human individual using a KOD Hot Start DNA Polymerase (EMD Millipore Corp., US) with primers of 5’-AA<u>GGTACC</u>TTACTCCTGGTGTAGTCGCATC -3’ and 5’-AAAA<u>CTCGAG</u>TGGCAGTGGTGGAAGCACAATA -3’, which were designed to introduce KpnI and XhoI restriction enzyme sites (underlined) at the 5’ and 3’ ends of the promoter sequence, respectively. These restriction sites were used in subcloning of the PCR product into pGL4.10[luc2] Firefly Luciferase Vector (Promega Corp., WI).

### Luciferase reporter assay

In 96-well white solid plates microplates, 4 × 10^4^ HepG2 cells were seeded and transiently transfected next day with 100 ng of plasmids which include 40 ng of *ZBTB20*-expression vector (pCMV6-*ZBTB20*) or empty expression vector (pCMV6-Entry Tagged Cloning Vector, OriGene Tech. Inc., MD) (Cat#: PS100001), 40 ng of pGL4.10-*AFP*, and 20 ng of *Renilla* Luciferase Control Reporter Vector as a transfection efficiency control (pRL-TK, Promega Corp., WI) using Lipofectamine™ 3000 Transfection Reagent (Thermo Fisher Scientific Inc., CA) according to the manufacturer’s instructions. Opti-MEM®| Reduced-Serum Medium (Gibco™) was replaced by Eagle’s Minimum Essential Medium (EMEM) (ATCC^®^) with 10% fetal bovine serum after 4 hours of transfection. Luciferase activity was measured using the Dual-Glo^®^ Luciferase Assay System (Promega Corp., WI) after 27 hours of transfection with Mithras LB940, Multimode Microplate Reader (Berthold Tech. GmbH & Co., Germany). All experiments were performed with four trials, with four replicates in each trial.

### Clinical Studies

Clinical and Neurological evaluations of the members of family PKST077 were performed at Allama Iqbal Medical Research Institute, Lahore, Pakistan and at the National Human Genome Research Institute, NIH. Brain MRI studies were available from individuals IV-24, IV-26, and IV-27 (Figure 1A).

### 2.10. KI mouse model

We used the CRISPR/Cas9 genome editing technology [27] to generate a knock-in mouse model that mimics the mutation founded in the affected members of stuttering family PKST77. (http://www.informatics.jax.org/marker/MGI:1929213), several different crRNAs were designed to target the sequence near to the variant and minimize potential off-target sites. More details are available in the Supplementary material 2.4 (Suppl.2.4).

### Ultrasound mouse vocalization analysis

The ultrasonic vocalizations of mice carrying the equivalent of the human Ile719Val mutation and their wild type littermates were gathered as previously described [12].

## Data Availability

All data produced in the present study are available upon reasonable request to the authors.

## Abbreviations used

NIDCD: National Institute on Deafness and Other Communication Disorders
ZBTB20: Zinc finger and BTB domain containing, 20
NHLBI: National Heart Lung and Blood Institute
CED: Communications Exome Database
AFP: Alpha feto protein
I_κ_B_α_: Inhibitor of kB alpha
GNPTAB: N-acetylglucosamine-1-phosphate transferase subunits alpha and beta
CRISPR-cas: Clustered Regularly Interspaced Short Palindromic Repeats-Crispr-associated protein 9
IRB: Institutional Review Board
NINDS: National Institute of Neurological Diseases and Stroke
PKSTR: Pakistani stuttering affected individuals, unrelated
PKNR: Pakistani normally fluent controls, unrelated
STCR: Cameroonian stuttering affected individuals, unrelated
RC: Cameroonian normal control individuals, unrelated
BRMII: Brazilian stuttering affected individuals, unrelated
BRCO: Brazilian normally fluent control individuals, unrelated
EMEM: Eagle’s Minimum Essential Medium
KI: Knock-in

## Declarations

### Ethical approval and consent to participate

Subjects were enrolled with written informed consent under NIH Institutional Review Board-approved Consent # 97-DC-0057 entitled Genetic Studies of Stuttering.

### Consent for publication

Subjects consented to publication without identifying information under NIH Institutional Review Board-approved Consent # 97-DC-0057 entitled Genetic Studies of Stuttering.

### Availability of Supporting data

Supporting data not presented in the Supplementary materials is available from the communicating author at

### Competing Interests

The authors declare no competing interests.

### Funding

This work was supported in part by the National Institute on Deafness and Other Communication Disorders (NIDCD) Intramural grant # Z01-DC-000047-19, the NIDCD Genomic and Computational Biology Core # Z1C-000086,

### Author contributions

CFD performed DNA sequencing, and linkage and bioinformatic analyses, MHR ascertained and sampled subjects and performed linkage analyses and DNA sequencing, TUH performed luciferase reporter studies, TB performed mouse ultrasonic vocalization studies, PS and GS evaluated MRI and other clinical data, SR ascertained subjects and supervised research in Pakistan, RM oversaw whole exome and whole genome sequencing, and DD conceived of the study, obtained funding, oversaw human subjects protection, and supervised all aspects of research. All authors contributed to writing and reviewing the manuscript.

## Acknowledgements

This work used the high performance computational capabilities of the Biowulf Linux Cluster at the National Institutes of Health, Bethesda, MD (http://biowulf.nih.gov). We thank Dr. Timothy Holy of Washington University St. Louis for support of TB, and The Stuttering Foundation, the National Stuttering Association, and Hollins Communications Research Institute, Roanoke, VA for assistance with subject recruitment.

## Competing Interests

The authors have no relevant financial or non-financial interests to disclose.

## Data Availability

The datasets generated during and/or analysed during the current study are available from the corresponding author on reasonable request.

## Notes

### Competing Interest Statement

Dennis Drayna serves on the Board of Directors of The Stuttering Foundation, a non-profit organization.

### Author Declarations

Human subjects, including individuals from a consanguineous Pakistani family (PKST77) with the inclusion diagnosis of non-syndromic persistent stuttering were enrolled with written informed consent under National Institutes of Health protocol 97-DC-0057 (Genetic studies of stuttering), also reviewed and approved by the IRB of the National Centre of Excellence in Molecular Biology, University of the Punjab.

