## Supplemental 1.1 for "Mutations in *ZBTB20* in individuals with persistent stuttering"

| **Reference** | **Variant** | | | **Protein domain** | **Origin** |
| --- | --- | --- | --- | --- | --- |
|  | **Exon** | **c.DNA** | **aa change** |  |  |
| Stellacci et al., 2018 | 4 | c.1024delC | p.(Gln342Serfs*42) | Protein lacking the entire C-terminal ZnF domain - DNA binding capacity disruption | De novo - heterozygote |
| Cleaver et al., 2019 |  | c.1749C>G | p.(Cys583Trp) | ZnF I | De novo - heterozygote |
| Cordeddu et al., 2014 |  | c.1768A>C | p.(Lys590Gln) |  | De novo - heterozygote |
| Cordeddu et al., 2014 |  | c.1771C>G | p.(Gln591Glu) |  | De novo - heterozygote |
| Cordeddu et al., 2014 |  | c.1787A>G | p.(His596Arg) |  | De novo - heterozygote |
| Casertano et al., 2016 |  | c.1787A>G | p.(His596Arg) |  | De novo - heterozygote |
| Grímsdóttir et al., 2019 |  | c.1800C>G | p.(His600Gln) |  | De novo - heterozygote |
| Cordeddu et al., 2014 |  | c.1802C>T | p.(Thr601Ile) |  | De novo - heterozygote |
| Cordeddu et al., 2014 | 5 | c.1805G>C | p.(Gly602Ala) |  | De novo - heterozygote |
| Cordeddu et al., 2014 |  | c.1811A>C | p.(Lys604Thr) |  | De novo - heterozygote |
| Ferreira et al., 2019 |  | c.1822T>C | p.(Cys608Arg) | ZnF II | na - heterozygote |
| Alby et al., 2018 |  | c.1832G>A | p.(Cys611Tyr) |  | De novo - heterozygote |
| Alby et al., 2018 |  | c.1837C>T | p.(Arg613Cys) |  | De novo - heterozygote |
| Mattioli et al., 2016 |  | c.1847C>T | p.(Ser616Phe) |  | De novo - heterozygote |
| Cleaver et al., 2019 |  | c.1850T>C | p.(Leu617Ser) |  | De novo - heterozygote |
| Cordeddu et al., 2014 |  | c.1861C>T | p.(Leu621Phe) |  | De novo - heterozygote |
| Casertano et al., 2016 |  | c.1869G>C | p.(Lys623Asn) |  | De novo - heterozygote |
| Ferreira et al., 2019 |  | c.1873A>G | p.(Met625Val) |  | na - heterozygote |
| Cordeddu et al., 2014 |  | c.1876G>A | p.(Val626Met) |  | na - heterozygote |
| Cleaver et al., 2019 |  | c.1879A>G | p.(Thr627Ala) |  | De novo - heterozygote |
| Alby et al., 2018 |  | c.1906T>C | p.(Cys636Arg) | ZnF III | De novo - heterozygote |
| Stellacci et al., 2018 |  | c.1931C>T | p.(Thr644Ile) |  | De novo - heterozygote |
| Cleaver et al., 2019 |  | c.1943C>T | p.(Ser648Phe) |  | De novo mosaic - heterozygote |
| Cleaver et al., 2019 |  | c.1967A>G | p.(His656Arg) |  | De novo - heterozygote |
| Mattioli et al., 2016 |  | c.2221G>A | p.(Gly741Arg) | ZnF V | De novo - heterozygote |

**Suppl. 1.1** Genetic variants reported in Primrose syndrome.
