## Supplemental 1.2 for "Mutations in *ZBTB20* in individuals with persistent stuttering"

| **1.2References** | **HGVS variants** | | **Reported phenotype** |
| --- | --- | --- | --- |
|  | **c.DNA** | **aa change** |  |
| Jones et al., 2018 | c.137C>G | p.(Pro46Arg) | Autism spectrum disorder |
| Alby et al., 2018; Kosmicki et al., 2017; McRae et al., 2017 | c.505G>C | p.(Glu169Gln) | Primrose syndrome |
| Alby et al., 2018; Kosmicki et al., 2017 | c.1020C>G | p.(Tyr340*) |  |
|  | c.1723A>T | p.(Lys575*) |  |
| Cleaver et al., 2018 | c.1749C>G | p.(Cys583Trp) |  |
| Cordeddu et al., 2014 | c.1768A>C | p.(Lys590Gln) |  |
|  | c.1771C>G | p.(Gln591Glu) |  |
| Cordeddu et al., 2014; Casertano et al., 2017 | c.1787A>G | p.(His596Arg) |  |
| Eldomery et al., 2017 | c.1786C>T | p.(His596Tyr) | Developmental delay/intellectual disability & other abnormalities |
| Grímsdóttir et al., 2019 | c.1800C>G | p.(His600Gln) | Primrose syndrome |
| Cordeddu et al., 2014 | c.1802C>T | p.(Thr601Ile) |  |
|  | c.1805G>C | p.(Gly602Ala) |  |
| Cordeddu et al., 2014; Farwell Hagman et al., 2017 | c.1811A>C | p.(Lys604Thr) | Multiple congenital anomalies |
| Martínez et al., 2017 | c.1832G>C | p.(Cys611Ser) | Primrose syndrome |
| Alby et al., 2018 | c.1832G>A | p.(Cys611Tyr) |  |
|  | c.1837C>T | p.(Arg613Cys) |  |
| Mattioli et al., 2016 | c.1847C>T | p.(Ser616Phe) |  |
| Cleaver et al., 2018 | c.1850T>C | p.(Leu617Ser) |  |
| Cordeddu et al., 2014 | c.1861C>T | p.(Leu621Phe) |  |
| Casertano et al., 2017 | c.1869G>C | p.(Lys623Asp) |  |
| Cordeddu et al., 2014 | c.1876G>A | p.(Val626Met) |  |
| Cleaver et al., 2018 | c.1879A>G | p.(Thr627Ala) |  |
| Alby et al., 2018 | c.1906T>C | p.(Cys636Arg) |  |

**Suppl.1.2** - *ZBTB20* – Variants reported in HGMD® Professional 2019.2

| c.1931C>T | p.T644I | Primrose syndrome | Stellacci et al., 2018 |
| --- | --- | --- | --- |
| c.1943C>T | p.S648F |  | Cleaver et al., 2018 |
| c.1967A>G | p.H656R |  |  |
| c.2221G>A | p.G741R |  | Mattioli et al., 2016 |
| c.1024delC | p.(Gln342Serfs*42) |  | Stellacci et al., 2018 |
| Gross deletions | | Developmental delay & autism | Rasmussen et al., 2014 |
|  |  | Hypotonia and language and motor delays | Shuvarikov et al., 2013 |
| Balanced 46,XY,t(3;8)(q13.31;q22.1) | | Developmental delay, attention-defecit hyperactivity disorder, Tourette's & autistic traits | Rasmussen et al., 2014 |
| Gross deletions | | Autism spectrum disorder | Wisniowiecka-Kowalnik et al., 2013 |
