## Supplemental 1.3 for "Mutations in *ZBTB20* in individuals with persistent stuttering"

| **Variant** | | | | **Stuttering Exome Cohort** | | | | | | | | | | | |
| --- | --- | --- | --- | --- | --- | --- | --- | --- | --- | --- | --- | --- | --- | --- | --- |
|  |  |  |  | **N = 1,941 (unrelated individuals)** | | | | | | | | | | | |
|  |  |  |  | **N = 1,074** | | | | |  | **N = 867** | | | | |  |
| **Exon** | **cDNA**  **position** | **aa**  **change** | **rsID** | **EurAm-st** | **Pk-st** | **Br-st** | **Cam-st** | | **All-st** | **EurAm-co** | | **Pk-co** | **Br-co** | **Cam-co** | **All-co** |
|  |  |  |  | **N=739** | **N=72** | **N=158** | **N=105** | |  | **N=460** | | **N=96** | **N=218** | **N=93** |  |
| 3 | c.52G>A | p.(Lys18Glu) | rs941801583 | 0 | 0 | 0 | 0 | | **0** | 0 | | 0 | 1 | 0 | **1** |
|  | c.154A>G | p.(Thr52Ala) | rs72950611 | 2 | 0 | 8 | 2 | | **12** | 0 | | 0 | 0 | 4 | **4** |
|  | c.178G>A | p.(Ala60Thr) | rs1484611510 | 0 | 0 | 0 | 0 | | **0** | 1 | | 0 | 0 | 0 | **1** |
|  | c.188G>A | p.(Gly63Glu) | rs182068139 | 0 | 0 | 1 | 0 | | **1** | 0 | | 0 | 0 | 0 | **0** |
| 4 | c.552A>G | p.(Thr184Thr) | rs na | 0 | 0 | 1 | 0 | | **1** | 0 | | 0 | 0 | 0 | **0** |
|  | c.569C>T | p.(Thr289Met) | rs1259818223 | 0 | 1 | 0 | 0 | | **1** | 0 | | 0 | 0 | 0 | **0** |
|  | c.735G>C | p.(Ser245Ser) | rs148117897 | 4 | 0 | 0 | 0 | | **4** | 0 | | 0 | 0 | 0 | **0** |
|  | c.750C>T | p.(Cys250Cys) | rs1230898999 | 1 | 0 | 0 | 0 | | **1** | 0 | | 0 | 0 | 0 | **0** |
|  | c.1067G>A | p.(Cys356Tyr) | rs na | 0 | 0 | 0 | 0 | | **0** | 1 | | 0 | 0 | 0 | **1** |
|  | c.1164C>G | p.(Asp388Glu) | rs144663365 | 6 | 1 | 0 | 0 | | **7** | 2 | | 1 | 3 | 0 | **6** |
|  | c.1250C>G | p.(Ala417Gly) | rs149352178 | 4 | 0 | 0 | 0 | | **4** | 4 | | 0 | 2 | 0 | **6** |
|  | c.1318G>A | p.(Glu440Lys) | rs138924453 | 18 | 2 | 3 | 0 | | **23** | 7 | | 2 | 4 | 0 | **13** |
|  | c.1404G>A | p.(Gln468Gln) | rs140765510 | 4 | 0 | 0 | 0 | | **4** | 3 | | 0 | 2 | 0 | **5** |
|  | c.1551G>A | p.(Ala517Ala) | rs747213174 | 2 | 0 | 0 | 0 | | **2** | 0 | | 0 | 0 | 0 | **0** |
|  | c.1557T>G | p.(Ser519Arg) | rs na | 1 | 0 | 0 | 0 | | **1** | 0 | | 0 | 0 | 0 | **0** |
|  | c.1638T>G | p.(Gly546Gly) | rs777884900 | 0 | 0 | 0 | 0 | | **0** | 1 | | 0 | 0 | 0 | **1** |
| 5 | c.1935G>A | p.(Gln645Gln) | rs374751714 | 0 | 1 | 0 | 0 | | **1** | 0 | | 0 | 0 | 0 | **0** |
|  | c.1950C>T | p.(Asn650Asn) | rs144019743 | 2 | 0 | 0 | 0 | | **2** | 2 | | 0 | 0 | 0 | **2** |
|  | c.2078C>T | p.(Ala679Val) | rs150974347 | 0 | 0 | 1 | 0 | | **1** | 0 | | 0 | 0 | 0 | **0** |
|  | c.2099G>A | p.(Arg700His) | rs760379885 | 0 | 0 | 0 | 0 | | **0** | 1 | | 0 | 0 | 0 | **1** |
|  | c.2155G>A | p.(Val719Ile) | rs779910215* | 0 | 0 | 0 | 0 | | **0** | 0 | | 0 | 0 | 0 | **0** |
|  | c.2199C>T | p.(His713His) | rs142627077 | 1 | 0 | 0 | 0 | | **1** | 0 | | 0 | 0 | 0 | **0** |
|  |  |  | **Total** | **45** | **5** | **14** | **2** | | **66** | **22** | | **3** | **12** | **4** | **41** |
|  |  | **Chi-square** | 0.9175 |  | p = 0.3381 | | | | |  | |  |  |  |  |
|  |  |  | 1.3234 | |  | p = 0.25 | | | | | |  |  |  |  |
|  |  |  | 1.6030 | | |  | | p = 0.205 | | | | |  |  |  |
|  |  |  | 0.9637 | | | |  | | p = 0.3262 | | | | |  |  |
|  |  |  | 1.8474 | | | | | |  | | p = 0.1741 | | | |  |
| Number of mutation carriers identified in stuttering case and controls separated by subpopulation and Chi-square P values comparing frequency in cases vs. controls. *Legend*: Variants determined by Whole-exome sequencing and validated by dideoxy- Sanger DNA sequencing in case and control populations. EurAm-st, European American stuttering cases, EurAm-co, European American controls; Pk-st, Pakistani stuttering cases, Pk-co, Pakistani controls; Br-st, Brazilian stuttering cases, Br-co, Brazilian controls; Cam-st, Cameroonian stuttering cases, Cam-Co, Cameroonian controls; All-st, all stuttering cases combined, All-co, all controls combined; rs na, rs not annotated; * PKST77 variant | | | | | | | | | | | | | | | |

**Supplement Table 1.3** Distribution of rare variants in *ZBTB20* (NM_001164342) in stuttering cases and controls subpopulation
