## Supplemental 2.2 for "Mutations in *ZBTB20* in individuals with persistent stuttering"

**Suppl. 2**  bcbio-nextgen pipeline configuration, tools base template.

| **Tools** | **Version used** | **Reference link (last releases)** | **Packages and bundles** |
| --- | --- | --- | --- |
| bamtools | 2.4.0 | [https://github.com/pezmaster31/bamtools](about:blank) | hg38-noalt-sequence (seq 0.1)  ([https://anaconda.org/ggd-alpha/hg38-noalt-sequence](about:blank))  hg38-noalt-bowtie2 v.0.1  ([https://anaconda.org/ggd-alpha/hg38-noalt-bowtie2](about:blank))  hg38-noalt-bwa v.0.1  ([https://anaconda.org/ggd-alpha/hg38-noalt-bwa](about:blank))  hg38-noalt-coverage, release date: 2015-11-17  ([https://anaconda.org/ggd-alpha/hg38-noalt-coverage](about:blank))  hg38-noalt-dbsnp, 147, release date: 2016-04-07  ([https://anaconda.org/ggd-alpha/hg38-noalt-dbsnp](about:blank))  hg38-noalt-hapmap_snps (v2.8), release date: 2015-05-22  ([https://anaconda.org/ggd-alpha/hg38-noalt-hapmap_snps/files](about:blank))  hg38-noalt-1000g_omni_snps (v2.8), release date: 2015-05-22  ([https://anaconda.org/ggd-alpha/hg38-noalt-1000g_omni_snps](about:blank))  hg38-noalt-1000g_snps, release date: 2015-05-22  (v2.8) ([https://anaconda.org/ggd-alpha/hg38-noalt-1000g_snps](about:blank))  hg38-noalt-mills_indels (v2.8), release date: 2015-05-22  ([https://anaconda.org/ggd-alpha/hg38-noalt-mills_indels](about:blank))  hg38-noalt-1000g_indels (v2.8), release date: 2015-05-22  ([https://anaconda.org/ggd-alpha/hg38-noalt-1000g_indels](about:blank))  hg38-noalt-clinvar, release date: 2015-03-30-2  ([https://anaconda.org/ggd-alpha/hg38-noalt-clinvar](about:blank))  hg38-noalt-transcripts, release date: 2015-11-22  ([https://anaconda.org/ggd-alpha/hg38-noalt-transcripts](about:blank)) |
| bcbio-nextgen | 0.9.9 | [https://github.com/bcbio/bcbio-nextgen](about:blank) |  |
| bcbio-variation | 0.2.6 | [https://github.com/chapmanb/bcbio.variation](about:blank) |  |
| bcftools | 1.3 | [https://samtools.github.io/bcftools/bcftools.html](about:blank) |  |
| bedtools | 2.24.0 | [https://github.com/arq5x/bedtools2/releases](about:blank) |  |
| biobambam | 2.0.44 | [https://github.com/gt1/biobambam2](about:blank) |  |
| bioconductor-bubbletree | 2.1.5 | [https://bioconductor.org/packages/3.9/bioc/html/BubbleTree.html](about:blank) |  |
| bowtie2 | 2.2.8 | [https://sourceforge.net/projects/bowtie-bio/files/bowtie2/2.2.8/](about:blank) |  |
| bwa | 0.7.15 | [http://bio-bwa.sourceforge.net/](about:blank) |  |
| chanjo | na | [https://github.com/Clinical-Genomics/chanjo](about:blank) |  |
| CNVkit | 0.7.11 | [https://pypi.org/project/CNVkit/0.7.11/](about:blank) |  |
| cufflinks | 2.2.1 | [https://github.com/cole-trapnell-lab/cufflinks](about:blank) |  |
| cutadapt | 1.1 | [https://cutadapt.readthedocs.io/en/v2.5/index.html](about:blank) |  |
| FastQC | 0.11.5 | [https://www.bioinformatics.babraham.ac.uk/projects/fastqc/](about:blank) |  |
| featureCounts | 1.4.4 | [http://bioinf.wehi.edu.au/featureCounts/](about:blank) |  |
| freebayes | 1.0.2.29 | [https://github.com/ekg/freebayes](about:blank) |  |
| GATK | 3.6-0 | [https://software.broadinstitute.org/gatk/documentation/tooldocs/3.6-0/](about:blank) |  |
| gatk-framework | 3.5.21 | [https://bioconda.github.io/recipes/gatk-framework/README.html](about:blank) |  |
| gemini | 0.18.3 | [https://anaconda.org/bioconda/gemini/files](about:blank) |  |
| grabix | 0.1.6 | [https://anaconda.org/bioconda/grabix/files](about:blank) |  |
| hisat2 | 2.0.4 | [http://daehwankimlab.github.io/hisat2/](about:blank) |  |
| htseq | 0.6.1 | [https://github.com/simon-anders/htseq](about:blank) |  |
| lumpy-sv | 0.2.12 | [https://github.com/arq5x/lumpy-sv](about:blank) |  |
| manta | 0.29.6 | [https://github.com/Illumina/manta](about:blank) |  |
| metasv | 0.4.0 | [https://github.com/bioinform/metasv](about:blank) |  |

| **Tools** | **Version used** | **Reference link (last releases)** |
| --- | --- | --- |
| mutect | 1.1.5 | [https://github.com/broadinstitute/mutect/releases](about:blank) |
| novoalign | 3.04.04 | [http://www.novocraft.com/products/novoalign/](about:blank) |
| novosort | 3.00.02 | [http://www.novocraft.com/products/novosort/](about:blank) |
| oncofuse | 1.1.0 | [https://github.com/mikessh/oncofuse](about:blank) |
| phylowgs | na | [https://github.com/morrislab/phylowgs](about:blank) |
| Picard | 1.141 | [https://broadinstitute.github.io/picard/](about:blank) |
| Platypus-variant | 0.8.1 | [https://github.com/andyrimmer/Platypus](about:blank) |
| qualimap | 2.2 | [http://qualimap.bioinfo.cipf.es/doc_html/intro.html](about:blank) |
| STAR | na | [https://github.com/alexdobin/STAR](about:blank) |
| rtg-tools | 3.6 | [https://github.com/RealTimeGenomics/rtg-tools](about:blank) |
| Sailfish | 0.10.1 | [http://www.cs.cmu.edu/~ckingsf/software/sailfish/](about:blank) |
| Salmon | 0.6.0 | [https://salmon.readthedocs.io/en/latest/salmon.html](about:blank) |
| sambamba | 0.6.3 | [https://github.com/biod/sambamba](about:blank) |
| samblaster | 0.1.22 | [https://github.com/GregoryFaust/samblaster](about:blank) |
| samtools | 1.3.1 | [https://sourceforge.net/projects/samtools/files/samtools/1.3.1/](about:blank) |
| scalpel | 0.5.1 | [https://sourceforge.net/projects/scalpel/](about:blank) |
| SnpEff | 4.2 | [http://snpeff.sourceforge.net/](about:blank) |
| VarDict | na | [https://github.com/AstraZeneca-NGS/VarDict](about:blank) |
| VarDict-Java | 1.4.6 | [https://github.com/AstraZeneca-NGS/VarDictJava](about:blank) |
| Variant-Effect-Predictor | na | [https://useast.ensembl.org/info/docs/tools/vep/index.html](about:blank) |
| Varscan | 2.4.1 | [http://varscan.sourceforge.net/](about:blank) |
| vcflib | 1.0.0_rc1 | [https://github.com/vcflib/vcflib](about:blank) |
| Vt | na | [https://genome.sph.umich.edu/wiki/Vt](about:blank) |
| wham | 1.7.0.162 | [https://github.com/zeeev/wham](about:blank) |

**Suppl.** bcbio-nextgen pipeline configuration, base template (cont.)
