## Supplemental 2.3 for "Mutations in *ZBTB20* in individuals with persistent stuttering"

| **Suppl.** *ZBTB20* (NM_ 001164342) Sanger sequence: PCR and Sequencing primers list | | |
| --- | --- | --- |
| PCR and Sequencing Primers (5'-3') | | |
| Region | Oligo ID | Sequence |
| Exon 01 | E1F*_pcr-seq_* | GAGGAAGTGCACAATGATTG |
|  | E1R*_seq_* | GAATAAGCCCTGTCATAATTAG |
|  | E1F*_seq_* | CAGGAGGAAGGTTCTGGTCG |
|  | E1R*_pcr-seq_* | TCAATGCCCTTGTTTCCTTC |
| Exon 02 and 03 | E2-3F*_pcr-seq_* | GAAACCTCCCATCACTACCCC |
|  | E2-3F*_seq_* | CATGAAAGCATATGGGCCAAG |
|  | E2-3R*_seq_* | CTTGGATAATGTCAGCCATACG |
|  | E2-3R*_pcr-seq_* | TGGTGGCACATTACTTTTGGC |
| Exon 04 | E4.1F*_pcr-seq_* | TTCCCTCAGGTCATCGTACCTA |
|  | E4.1R*_seq_* | CTGTTTTGATCTGCAGGATGC |
|  | E4.1F*_seq_* | GCATCCACGGGAGCATGCTGC |
|  | E4.1R*_pcr-seq_* | GATGTTGCCCACTAGGGTCTG |
|  | E4.2F*_pcr-seq_* | AGAGCAGCGACACGGAGT |
|  | E4.2R*_seq_* | CATCCTCAGGTTGCTGGTGAG |
|  | E4.2F*_seq_* | AGAAACAGGTGCTTCCTCTCC |
|  | E4.2R*_pcr-seq_* | TGCTGCCCGTAGTAGTCGTA |
| Exon 05 | E5F*_pcr-seq_* | TTGTTGTCTTGTTTTTCCTGTC |
|  | E5R*_seq_* | TGTTTTGTTCATAAGAAAGAGAG |
|  | E5F*_seq_* | GTTCTCTCACAAGACCCTCCTGGA |
|  | E5R*_pcr-seq_* | ACTGGTTACTGTGAAGTCCAAGC |
| Note: F=Forward; R = Reverse; pcr = fragment amplification; seq = BigDye reaction | | |
