## Supplemental 2.4 for "Mutations in *ZBTB20* in individuals with persistent stuttering"

### *Zbtb20* V719I
### Knock In with CRISPR/Cas9

#### Founder Report

The Jackson Laboratory
*Leading the search for tomorrow's cures*

### Project Goal

- The Jackson Laboratory will introduce a point mutation in the *Zbtb20* gene changing the valine at position 719 to isoleucine.

<table>
<tr><th colspan="2">Requestor</th><th>Dennis Drayna, NIH - NIDCD, Bethesda</th></tr>
<tr><td rowspan="3">Gene</td><td>Name</td><td>Zbtb20 (zinc finger and BTB domain containing 20)</td></tr>
<tr><td>Location</td><td>Chr16:42875881-43642602 bp, + strand<br>http://www.informatics.jax.org/marker/MGI:1929213</td></tr>
<tr><td>Transcript ID</td><td>ENSMUST00000114691.7<br>http://www.ensembl.org</td></tr>
<tr><td colspan="2">Previously Described Phenotypes</td><td>Mice homozygous for a knock-out allele exhibit growth retardation, disrupted homeostasis, and premature death.<br>http://www.informatics.jax.org</td></tr>
<tr><td colspan="2">Model Desired</td><td>V719I</td></tr>
<tr><td colspan="2">Strain (JAX Registry #)</td><td>C57BL/6J (JR000664)</td></tr>
<tr><td rowspan="2">CRISPR</td><td>Guides</td><td>IDT0911_Zbtb20_V719_crRNA1: GTCAAACTTTGCTGGGCAGA<br>IDT0912_Zbtb20_V719_crRNA2: CGATTTGGTCAAACTTTGCT<br>IDT0913_Zbtb20_V719_crRNA3: TCGATTTGGTCAAACTTTGC</td></tr>
<tr><td>Donor DNA</td><td>10012_Zbtb20_V719I_donor 1 (Forward; for use with guide 1):<br>ACCCCTCCGGCAGGCACGCCCCCAGGTGCCCGCGCGGGTCCGCCAGGCGTGGTGGCCTGCACAGAG<br>GGGACCACTTACGTCTGCTCTATCTGCCCAGCAAAGTTTGACCAAATCGAGCAGTTCAACG<br><br>10012_Zbtb20_V719I_donor 2 (forward; for use with guide 2 or 3):<br>GGCAGGCACGCCCCCAGGTGCCCGCGCGGGTCCGCCAGGCGTGGTGGCCTGCACAGAGGGGACCAC<br>TTACGTCTGCTCCATCTGCCCAGCTAAGTTTGACCAAATCGAGCAGTTCAACGACCACATG</td></tr>
</table>

### Mouse *Zbtb20* transcripts

#### Gene: Zbtb20 ENSMUSG00000022708

| | |
|---|---|
| Description | zinc finger and BTB domain containing 20 [Source:MGI Symbol;Acc:MGI:1929213] |
| Gene Synonyms | 1300017A20Rik, 7330412A13Rik, A930017C21Rik, D16Wsu73e, HOF, Zfp288 |
| Location | Chromosome 16: 42,875,881-43,642,602 forward strand.<br>GRCm38:CM001009.2 |
| About this gene | This gene has 21 transcripts (splice variants), 130 orthologues, is a member of 1 Ensembl protein family and is associated with 42 phenotypes. |
| Transcripts | Hide transcript table |

Show/hide columns (1 hidden) Filter

| Name | Transcript ID | bp | Protein | Biotype | CCDS | UniProt | RefSeq | Flags |
|---|---|---|---|---|---|---|---|---|
| Zbtb20-201 | ENSMUST00000079441.12 | 3086 | 741aa | Protein coding | CCDS28178 | Q8K0L9 | NM_019778<br>NP_062752 | TSL:5 GENCODE basic APPRIS P3 |
| Zbtb20-202 | ENSMUST00000114690.7 | 2759 | 668aa | Protein coding | CCDS49851 | Q8K0L9 | - | TSL:1 GENCODE basic APPRIS ALT2 |
| Zbtb20-203 | ENSMUST00000114691.7 | 2891 | 668aa | Protein coding | CCDS49851 | Q8K0L9 | NM_181058<br>NP_851401 | TSL:1 GENCODE basic APPRIS ALT2 |
| Zbtb20-204 | ENSMUST00000114694.8 | 26901 | 741aa | Protein coding | CCDS28178 | Q8K0L9 | NM_001285805<br>NP_001272734 | TSL:5 GENCODE basic APPRIS P3 |
| Zbtb20-205 | ENSMUST00000114695.2 | 2977 | 741aa | Protein coding | CCDS28178 | Q8K0L9 | - | TSL:1 GENCODE basic APPRIS P3 |
| Zbtb20-206 | ENSMUST00000122875.7 | 736 | No protein | Processed transcript | - | - | - | TSL:3 |
| Zbtb20-207 | ENSMUST00000123047.7 | 439 | 3aa | Protein coding | - | - | - | CDS 3' incomplete TSL:3 |
| Zbtb20-208 | ENSMUST00000126100.7 | 463 | 3aa | Protein coding | - | - | - | CDS 3' incomplete TSL:5 |
| Zbtb20-209 | ENSMUST00000126354.7 | 563 | No protein | Retained intron | - | - | - | TSL:3 |
| Zbtb20-210 | ENSMUST00000131689.7 | 425 | No protein | Processed transcript | - | - | - | TSL:2 |
| Zbtb20-211 | ENSMUST00000132881.7 | 418 | No protein | Processed transcript | - | - | - | TSL:3 |
| Zbtb20-212 | ENSMUST00000134537.2 | 645 | No protein | Retained intron | - | - | - | TSL:3 |
| Zbtb20-213 | ENSMUST00000139491.7 | 2063 | No protein | Retained intron | - | - | - | TSL:1 |
| Zbtb20-214 | ENSMUST00000141941.7 | 414 | No protein | Processed transcript | - | - | - | TSL:3 |
| Zbtb20-215 | ENSMUST00000146708.7 | 821 | 75aa | Protein coding | - | E0CXK1 | - | CDS 3' incomplete TSL:3 |
| Zbtb20-216 | ENSMUST00000148775.7 | 590 | 60aa | Protein coding | - | E0CXT9 | - | CDS 3' incomplete TSL:3 |
| Zbtb20-217 | ENSMUST00000151244.7 | 754 | 63aa | Protein coding | - | E0CX28 | - | CDS 3' incomplete TSL:3 |
| Zbtb20-218 | ENSMUST00000151290.7 | 593 | No protein | Retained intron | - | - | - | TSL:3 |
| Zbtb20-219 | ENSMUST00000156367.7 | 787 | 131aa | Protein coding | - | E0CYJ9 | - | CDS 3' incomplete TSL:3 |
| Zbtb20-220 | ENSMUST00000156981.7 | 837 | 138aa | Protein coding | - | E0CYH3 | - | CDS 3' incomplete TSL:3 |
| Zbtb20-221 | ENSMUST00000162745.7 | 267 | No protein | Processed transcript | - | - | - | TSL:5 |

Project Goal: Introduce a point mutation in the *Zbtb20* gene (V719I).

www.ensembl.org

### Mouse *Zbtb20* transcripts

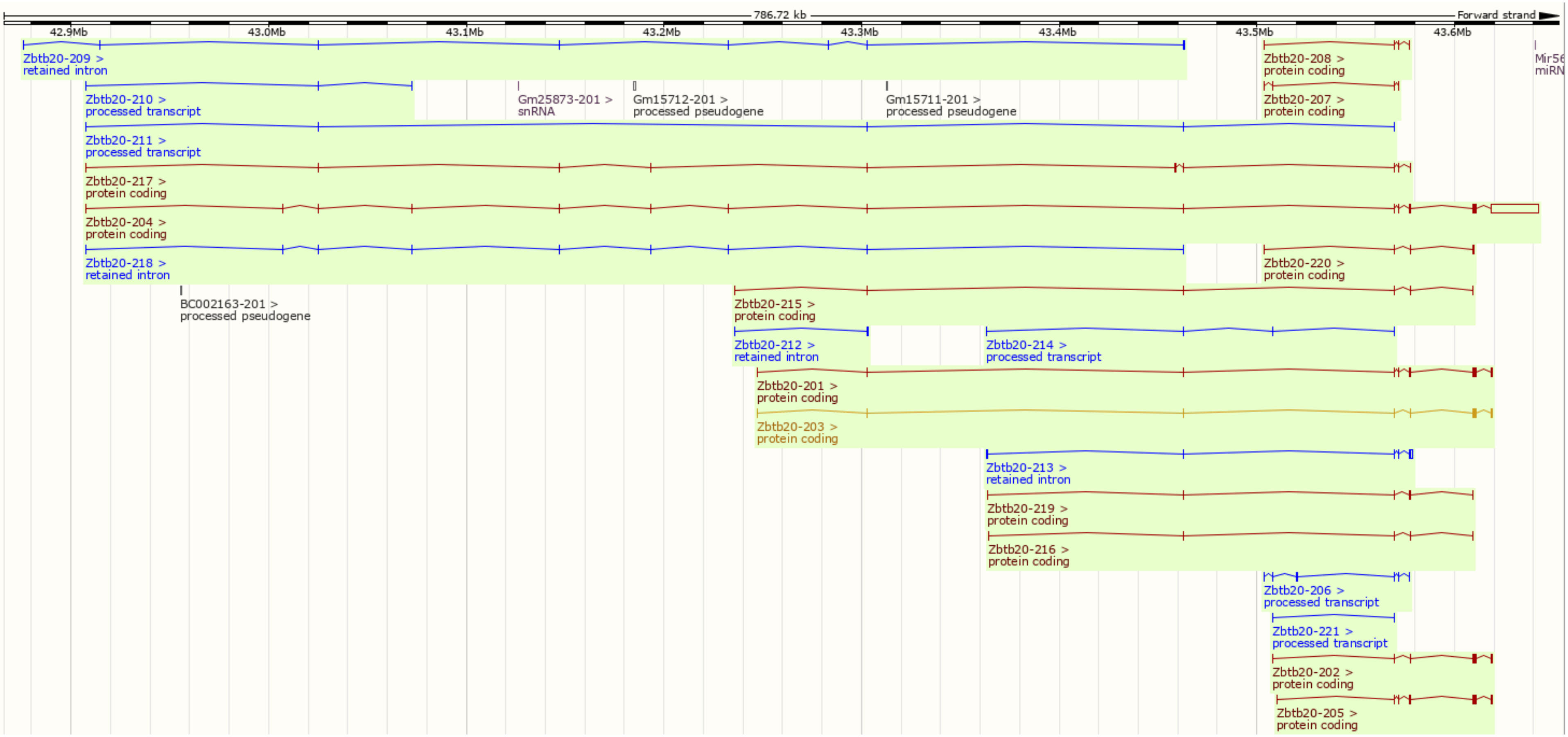


Project Goal: Introduce a point mutation in the *Zbtb20* gene (V719I). www.ensembl.org

### *Zbtb20* V719I- Targeting Strategy 1

WT allele

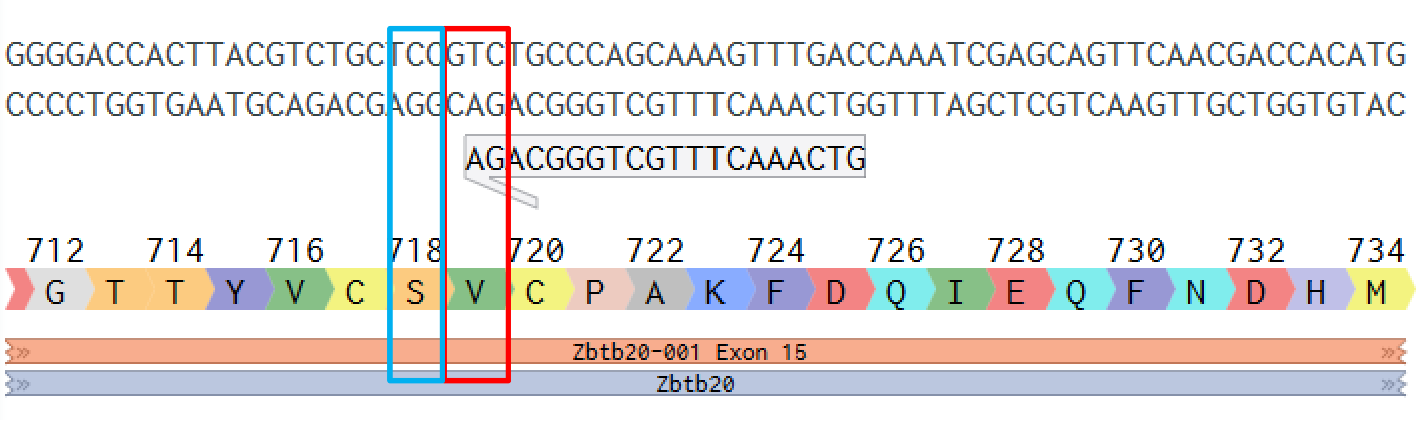


TCC>TCT

*Silent Mutation

V719I; GTC > ATC

KI allele

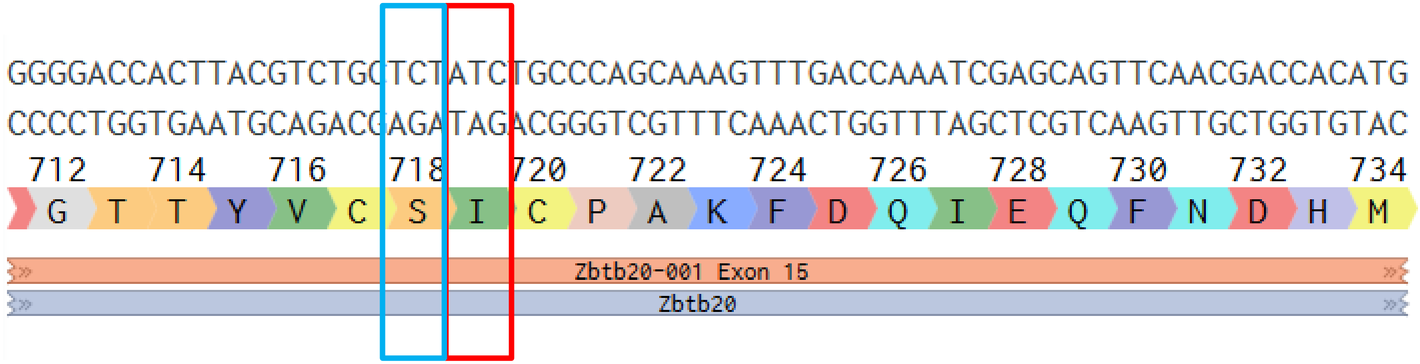


www.benchling.com

### *Zbtb20* V719I- Targeting Strategy 2

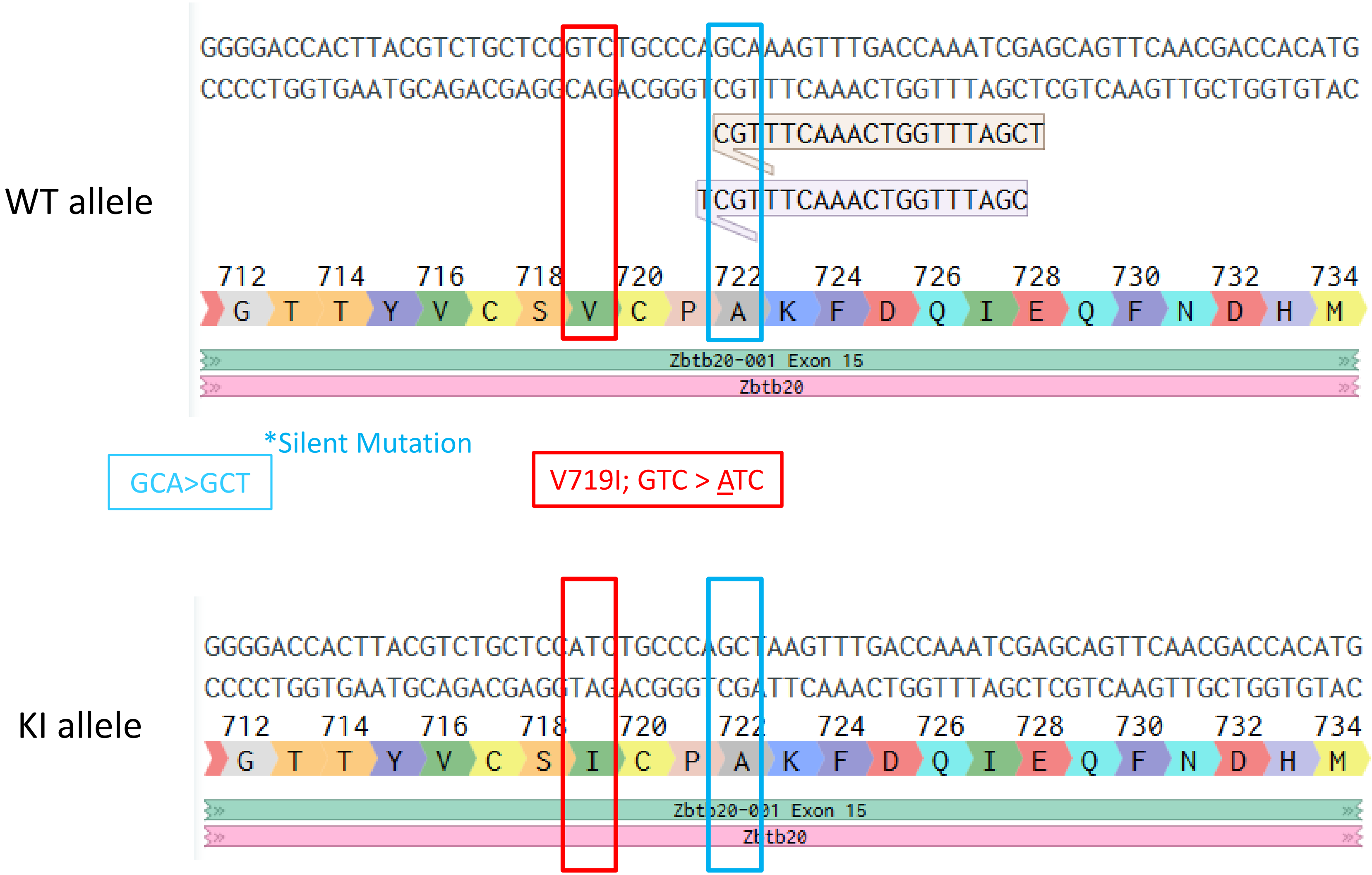


www.benchling.com

### *Zbtb20* V719I– Potential Off-Target Sites

Zbtb20_V719_crRNA1: GTCAAACTTTGCTGGGCAGA

| Sequence | PAM | Score | Gene | Locus |
|---|---|---|---|---|
| GTCAAACTTTGCTGGGCAGA | CGG | 100.0 | Zbtb20 (ENSMUSG00000022708) | chr16:-43618677 |
| TTTCAACTTTGCTGGGCAGA | CAG | 2.4 | | chr15:-44730934 |
| ATAGAACCTTGCTGGGCAGA | AAG | 1.5 | | chr9:-85801466 |
| CTCAATCTTTGCTGGGCAGC | TAG | 1.2 | | chr12:+65392930 |

Zbtb20_V719_crRNA2: CGATTTGGTCAAACTTTGCT

| Sequence | PAM | Score | Gene | Locus |
|---|---|---|---|---|
| CGATTTGGTCAAACTTTGCT | GGG | 100.0 | Zbtb20 (ENSMUSG00000022708) | chr16:-43618684 |
| TCATTTGGTTAAACTTTGCT | GAG | 2.7 | | chr7:+87971054 |
| TTAATTGGTCAAACTTTGCT | GGG | 2.4 | | chr14:+12226611 |
| CGGGTTGGCCAAACTTTGCT | TAG | 1.6 | | chr1:+191432251 |
| CTATGTGCTTAAACTTTGCT | GAG | 1.4 | | chr16:-88425655 |

Zbtb20_V719_crRNA3: TCGATTTGGTCAAACTTTGC

| Sequence | PAM | Score | Gene | Locus |
|---|---|---|---|---|
| TCGATTTGGTCAAACTTTGC | TGG | 100.0 | Zbtb20 (ENSMUSG00000022708) | chr16:-43618685 |
| TTTAATTGGTCAAACTTTGC | TGG | 2.4 | | chr14:+12226610 |
| TTAATGTGGTCAAACTTTGC | TAG | 1.5 | | chr9:-52623784 |
| TGGATTTGGACAAACTTTCC | AAG | 1.2 | | chr3:-104528297 |
| CAGACTGGGTCAAACTTTGC | CAG | 1.0 | | chr9:+110042760 |

Off-target scoring: Higher off-target editing risk when score is >2.0 with a canonical PAM (NGG) or >50.0 with a unlinked non-canonical PAM (NAG) or >20.0 with a linked non-canonical PAM in a coding region.

### *Zbtb20* V719I– Sequencing Strategy

KI allele

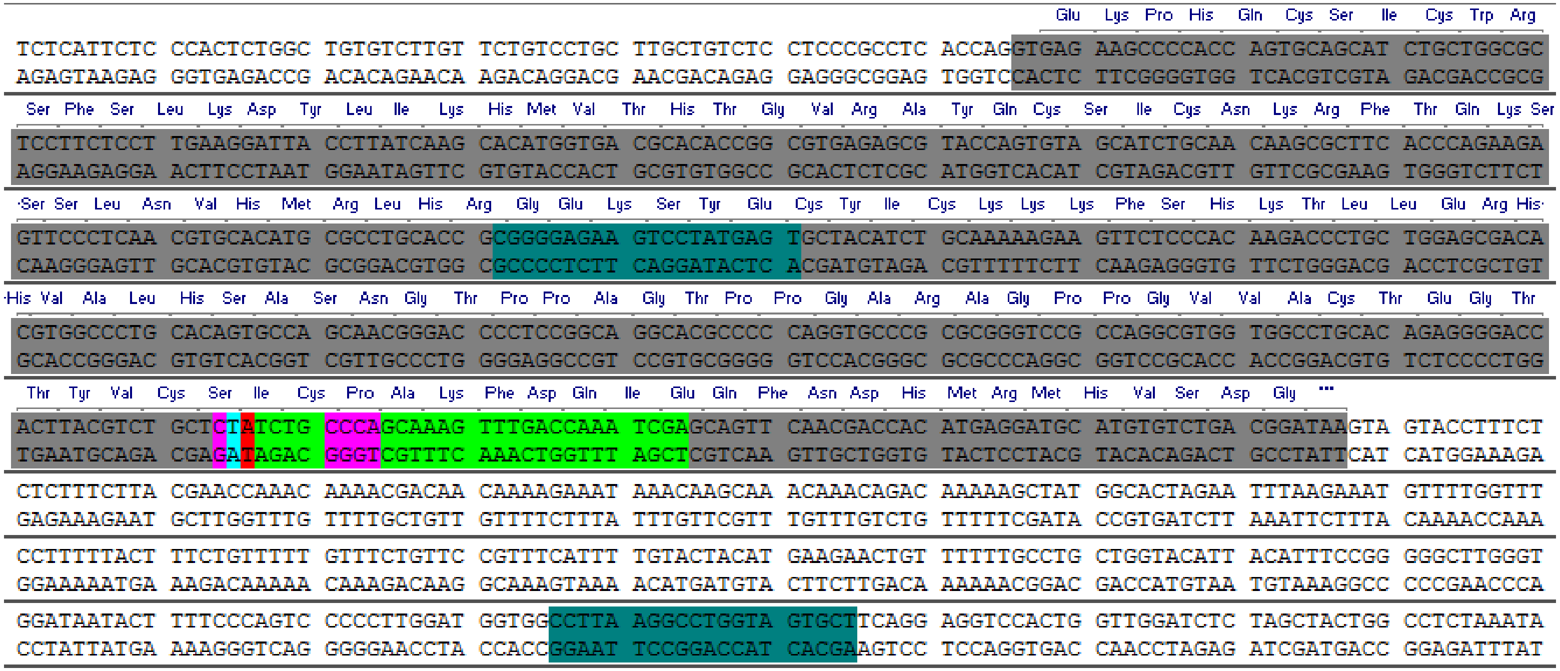


**#1: Product of length 524 (rating: 175)**

Contains region of the molecule from 632 to 1155

Tm: 78.7 C TaOpt: 55.1 C GC: 49.0

Sense Primer:

CGGGGAGAAGTCCTATGAGT

Similarity: 100.0%

Length: 20 Tm: 49.8 C GC: 55.0

dH: -152.0 kcal/mol dS: -394.5 cal/mol dG: -32.6 kcal/mol

Antisense Primer:

AGCACTACCAGGCCTTAAGG

Similarity: 100.0%

Length: 20 Tm: 50.5 C GC: 55.0

dH: -160.0 kcal/mol dS: -416.8 cal/mol dG: -34.0 kcal/mol

Tm Difference: 0.7

GC Difference: 0.0

V719I; GTC > <u>A</u>TC

TCC>TCT

Guide | PAM | Primer

**<u>PRIMERS</u>**

10014_Zbtb20_genoF: CGGGGAGAAGTCCTATGAGT

10015_Zbtb20_genoR: AGCACTACCAGGCCTTAAGG

Strain#: 408764

Request#: 552388

Animal ID: 4695- Female; 4698, 4726, 4727, 4728- Male

V719I KI

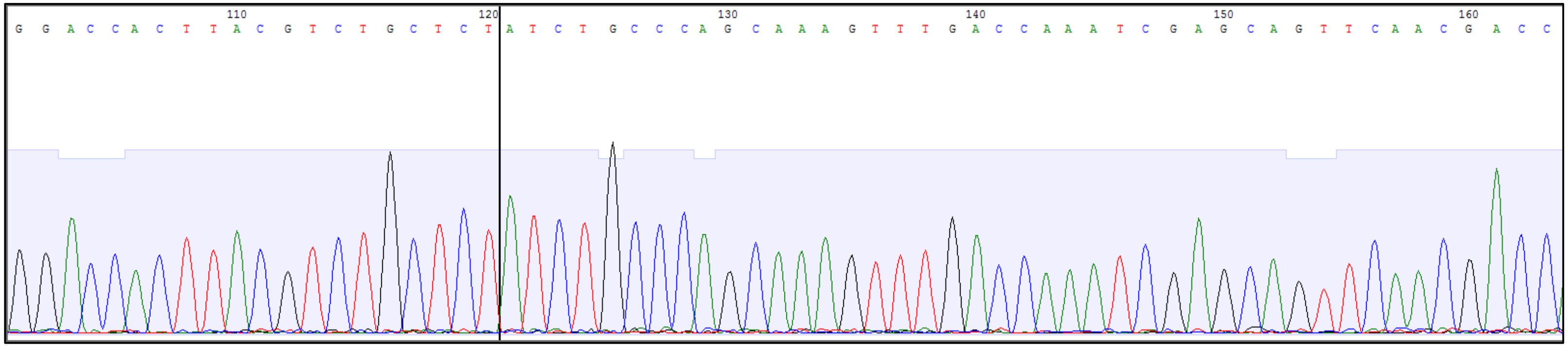


WT: GGACCACTTACGTCTGCTCCGTCTGCCCAGCAAAGTTTGACCAAATCGAGCAGTTCAACGAC

KI: GGACCACTTACGTCTGCTCTATCTGCCCAGCAAAGTTTGACCAAATCGAGCAGTTCAACGAC

TCC>TCT

V719I; GTC > ATC
