## Supplemental 3.2 for "Mutations in *ZBTB20* in individuals with persistent stuttering"

| **Suppl.** *ZBTB20* multiple sequence alignment in 99 vertebrates. Bolded and grey letter denote residue Valine 719 conserved on 95/99 vertebrates. | | | | | | | | |
| --- | --- | --- | --- | --- | --- | --- | --- | --- |
| **Assembly** | **Species name** | **Residue 719** | | **Assembly** | | **Species name** | **p.Val719Ile** | |
| hg38 | *Homo sapiens* | 709-TEGTTYVCS**V**CPAKFDQ-727 | | eleEdw1 | | *Elephantulus edwardii* | 709-TEGTTYVCS**V**CPAKFDQ-727 | |
| panTro4 | *Pan troglodytes* | 709-TEGTTYVCS**V**CPAKFDQ-727 | | triMan1 | | *Trichechus manatus latirostris* | 709-TEGTTYVCS**V**CPAKFDQ-727 | |
| gorGor3 | *Troglodytes gorilla* | 709-TEGTTYVCS**V**CPAKFDQ-727 | | chrAsi1 | | *Chrysochloris asiatica* | 709-TEGTTYVCS**V**CPAKFDQ-727 | |
| ponAbe2 | *Pongo pygmaeus abelii* | 709-TEGTTYVCS**V**CPAKFDQ-727 | | echTel2 | | *Echinops telfairi* | 709-TEGTTYVCS**V**CPAKFDQ-727 | |
| nomLeu3 | *Nomascus leucogenys* | 709-TEGTTYVCS**V**CPAKFDQ-727 | | oryAfe1 | | *Orycteropus afer* | 709-TEGTTYVCS**V**CPAKFDQ-727 | |
| rheMac3 | *Macaca mulatta* | 709-TEGTTYVCS**V**CPAKFDQ-727 | | dasNov3 | | *Dasypus novemcinctus* | 709-TEGTTYVCS**V**CPAKFDQ-727 | |
| macFas5 | *Macaca fascicularis* | 709-TEGTTYVCS**V**CPAKFDQ-727 | | monDom5 | | *Didelphis virginiana* | 709-TEGTTYVCS**V**CPAKFDQ-727 | |
| papAnu2 | *Papio hamadryas* | 709-TEGTTYVCS**V**CPAKFDQ-727 | | sarHar1 | | *Sarcophilus harrisii* | 709-TEGTTYVCS**V**CPAKFDQ-727 | |
| chlSab2 | *Chlorocebus sabaeus* | 709-TEGTTYVCS**V**CPAKFDQ-727 | | macEug2 | | *Macropus eugenii* | 709-------------------------------727 | |
| calJac3 | *Callithrix jacchus* | 709-TEGTTYVCS**V**CPAKFDQ-727 | | ornAna1 | | *Ornithorhynchus anatinus* | 709-TEGTTYVCS**V**CPAKFDQ-727 | |
| saiBol1 | *Saimiri boliviensis* | 709-TEGTTYVCS**V**CPAKFDQ-727 | | falChe1 | | *Falcon cherrug* | 709-TEGTTYVCS**V**CPAKFDQ-727 | |
| otoGar3 | *Otolemur garnettii* | 709-TEGTTYVCS**V**CPAKFDQ-727 | | falPer1 | | *Falco peregrinus* | 709-TEGTTYVCSVCPAKFDQ-727 | |
| tupChi1 | *Tupaia chinensis* | 709-TEGTTYVCS**V**CPAKFDQ-727 | | ficAlb2 | | *Ficedula albicollis* | 709-TEGTTYVCS**V**CPAKFDQ-727 | |
| speTri2 | *Spermophilus tridecemlineatus* | 709-TEGTTYVCS**V**CPAKFDQ-727 | | zonAlb1 | | *Zonotrichia albicollis* | 709-TEGTTYVCS**V**CPAKFDQ-727 | |
| jacJac1 | *Jaculus jaculus* | 709-TEGTTYVCS**V**CPAKFDQ-727 | | geoFor1 | | *Geospiza fortis* | 709-TEGTTYVCS**V**CPAKFDQ-727 | |
| micOch1 | *Microtus ochrogaster* | 709-TEGTTYVCS**V**CPAKFDQ-727 | | taeGut2 | | *Taeniopygia guttata* | 709-TEGTTYVCS**V**CPAKFDQ-727 | |
| criGri1 | *Cricetulus griseus* | 709-TEGTTYVCS**V**CPAKFDQ-727 | | pseHum1 | | *Pseudopodoces humilis* | 709-TEGTTYVCS**V**CPAKFDQ-727 | |
| mesAur1 | *Microtus ochrogaster* | 709-TEGTTYVCS**V**CPAKFDQ-727 | | melUnd1 | | *Melopsittacus undulatus* | 709-TEGTTYVCS**V**CPAKFDQ-727 | |
| mm10 | *Mus musculus* | 709-TEGTTYVCS**V**CPAKFDQ-727 | | amaVit1 | | *Amazona vittata* | 709-TEGTTYVCS**V**CPAKFDQ-727 | |
| rn6 | *Rattus norvegicus* | 709-TEGTTYVCS**V**CPAKFDQ-727 | | araMac1 | | *Ara macao* | 709-TEGTTYVCS**V**CPAKFDQ-727 | |
| hetGla2 | *Heterocephalus glaber* | 709-TEGTTYVCS**V**CPAKFDQ-727 | | colLiv1 | | *Columba livia* | 709-TEGTTYVCS**V**CPAKFDQ-727 | |
| cavPor3 | *Cavia porcellus* | 709-TEGTTYVCS**V**CPAKFDQ-727 | | anaPla1 | | *Anas platyrhynchos* | 709-TEGTTYVCS**V**CPAKFDQ-727 | |
| chiLan1 | *Chinchilla lanigera* | 709-TEGTTYVCS**V**CPAKFDQ-727 | | galGal4 | | *Gallus gallus* | 709-TEGTTYVCS**V**CPAKFDQ-727 | |
| octDeg1 | *Octodon degus* | 709-TEGTTYVCS**V**CPAKFDQ-727 | | melGal1 | | *Meleagris gallopavo* | 709-TEGTTYVCS**V**CPAKFDQ-727 | |
| oryCun2 | *Oryctolagus cuniculus* | 709-TEGTTYVCS**V**CPAKFDQ-727 | | allMis1 | | *Alligator mississippiensis* | 709-TEGTTYVCS**V**CPAKFDQ-727 | |
| ochPri3 | *Ochotona princeps* | 709-TEGTTYVCS**V**CPAKFDQ-727 | | cheMyd1 | | *Chelonia mydas* | 709-TEGTTYVCS**V**CPAKFDQ-727 | |
| susScr3 | *Sus scrofa* | 709-TEGTTYVCS**V**CPAKFDQ-727 | | chrPic2 | | *Chrysemys picta bellii* | 709-TEGTTYVCS**V**CPAKFDQ-727 | |
| vicPac2 | *Vicugna pacos* | 709-TEGTTYVCS**V**CPAKFDQ-727 | | pelSin1 | | *Pelodiscus sinensis* | 709-TEGTTYVCS**V**CPAKFDQ-727 | |
| **Suppl.** *ZBTB20* multiple sequence alignment in 99 vertebrates. Bolded and grey letter denote residue Valine 719 conserved on 95/99 vertebrates. *(cont.)* | | | | | | | | |
| **Assembly** | **Species name** | **Residue 719** | **Assembly** | | **Species name** | | | **p.Val719Ile** |
| camFer1 | *Camelus ferus* | 709-TEGTTYVCS**V**CPAKFDQ-727 | apaSpi1 | | *Apalone spinifera* | | | 709--EGTTYVCS**V**CPAKFDQ-727 |
| turTru2 | *Tursiops truncatus* | 709-TEGTTYVCS**V**CPAKFDQ-727 | anoCar2 | | *Anolis carolinensis* | | | 709-AEGTTYVCS**V**CPAKFDQ-727 |
| orcOrc1 | *Orcinus orca* | 709-TEGTTYVCS**V**CPAKFDQ-727 | xenTro7 | | *Xenopus tropicalis* | | | 709-AEGTTYVCS**V**CPTKFDQ-727 |
| panHod1 | *Pantholops hodgsonii* | 709-TEGTTYVCS**V**CPAKFDQ-727 | latCha1 | | *Latimeria chalumnae* | | | 709-TGTVSZACSFYPTCLLKN-727 |
| bosTau8 | *Bos taurus* | 709-TEGTTYVCS**V**CPAKFDQ-727 | tetNig2 | | *Tetraodon lineatus* | | | 709-QEGTTYVCS**V**CPAKFDQ-727 |
| oviAri3 | *Ovis aries* | 709-TEGTTYVCS**V**CPAKFDQ-727 | fr3 | | *Takifugu rubripes* | | | 709-QEGTTYVCS**V**CPAKFDQ-727 |
| capHir1 | *Capra hircus* | 709-TEGTTYVCS**V**CPAKFDQ-727 | takFla1 | | *Takifugu flavidus* | | | 709-QEGTTYVCS**V**CPAKFDQ-727 |
| cerSim1 | *Ceratotherium simum* | 709-TEGTTYVCS**V**CPAKFDQ-727 | oreNil2 | | *Oreochromis niloticus* | | | 709-QEGTTYVCS**V**CPAKFDQ-727 |
| felCat8 | *Felis catus* | 709-TEGTTYVCS**V**CPAKFDQ-727 | neoBri1 | | *Neolamprologus brichardi* | | | 709-QEGTTYVCS**V**CPAKFDQ-727 |
| canFam3 | *Canis lupus familiaris* | 709-TEGTTYVCS**V**CPAKFDQ-727 | hapBur1 | | *Haplochromis burtoni* | | | 709-QEGTTYVCS**V**CPAKFDQ-727 |
| musFur1 | *Mustela putorius furo* | 709-TEGTTYVCS**V**CPAKFDQ-727 | mayZeb1 | | *Maylandia zebra* | | | 709-QEGTTYVCS**V**CPAKFDQ-727 |
| ailMel1 | *Ailuropoda melanoleuca* | 709-TEGTTYVCS**V**CPAKFDQ-727 | punNye1 | | *Pundamilia nyererei* | | | 709-QEGTTYVCS**V**CPAKFDQ-727 |
| odoRosDiv1 | *Odobenus rosmarus divergens* | 709-TEGTTYVCS**V**CPAKFDQ-727 | oryLat2 | | *Oryzias latipes* | | | 709-QEGTTYVCS**V**CPAKFDQ-727 |
| lepWed1 | *Leptonychotes weddellii* | 709-TEGTTYVCS**V**CPAKFDQ-727 | xipMac1 | | *Xiphophorus maculatus* | | | 709-QEGTTYVCS**V**CPAKFDQ-727 |
| pteAle1 | *Pteropus alecto* | 709-TEGTTYVCS**V**CPAKFDQ-727 | gasAcu1 | | *Gasterosteus aculeatus* | | | 709-QEGTTYVCS**V**CPAKFDQ-727 |
| pteVam1 | *Pteropus vampyrus* | 709-TEGTTYVCS**V**CPAKFDQ-727 | gadMor1 | | *Gadus morhua* | | | 709----------------------AKFDQ-727 |
| myoDav1 | *Myotis davidii* | 709-TEGTTYVCS**V**CPAKFDQ-727 | danRer1 | | *Danio rerio* | | | 709-QEGTTYMCS**V**CPVKFDQ-727 |
| myoLuc2 | *Myotis lucifugus* | 709-TEGTTYVCS**V**CPAKFDQ-727 | astMex1 | | *Astyanax mexicanus* | | | 709-QEGTTYMCS**V**CPVKFDQ-727 |
| eptFus1 | *Eptesicus fuscus* | 709-TEGTTYVCS**V**CPAKFDQ-727 | lepOcu1 | | *Lepisosteus oculatus* | | | 709-QEGTTYVCS**V**CPAKFDQ-727 |
| eriEur2 | *Erinaceus europaeus* | 709-TEGTTYVCS**V**CPAKFDQ-727 | petMar2 | | *Petromyzon marinus* | | | 709-TGEKPYACD**V**C-A--HA-727 |
| sorAra2 | *Sorex araneus* | 709-TEGTTYVCS**V**CPAKFDQ-727 |  | |  | | |  |
| conCri1 | *Condylura cristata* | 709-TEGTTYVCS**V**CPAKFDQ-727 |  | |  | | |  |
| loxAfr3 | *Loxodonta africana* | 709-TEGTTYVCS**V**CPAKFDQ-727 |  | |  | | |  |
