## Supplemental 3.5 for "Mutations in *ZBTB20* in individuals with persistent stuttering"

| **Chr** | **Start** | **End** | **rsID** | **Ref** | **Alt** | **Region** | **Gene name** | **Type** | **Exon** | **c.DNA position** | **Amino Acid change** |
| --- | --- | --- | --- | --- | --- | --- | --- | --- | --- | --- | --- |
| 3 | 112113049 | 112113049 | rs191148598 | G | A | exonic | *C3orf52* | nonsynonymous | Exon 5 | c.553G>A | p.E185K |
| 3 | 112974799 | 112974801 | rs564728039 | AAG | - | exonic | *CD200R1* | nonframeshift deletion | Exon 1 | c.57_59del | p.19_20del |
| 3 | 112991054 | 112991054 | rs565265086 | G | A | exonic | *GTPBP8* | nonsynonymous | Exon 1 | c.55G>A | p.V19M |
| 3 | 114339076 | 114339076 | rs779910215 | C | T | exonic | *ZBTB20* | nonsynonymous | Exon 5 | c.2155G>A* | p.V719I |
| 3 | 119034641 | 119034641 | rs576681103 | G | C | UTR5 | *IGSF11* | - | - | - | - |
| 3 | 119147246 | 119147246 | rs554707851 | C | G | exonic | *C3orf30* | nonsynonymous | Exon 1 | c.1057C>G | p.Q353E |
| 3 | 119732456 | 119732456 | rs773076124 | A | G | exonic | *MAATS1* | nonsynonymous | Exon 9 | c.1181A>G | p.Y394C |
| 3 | 120348820 | 120348820 | rs201718706 | G | T | exonic | *LRRC58* | nonsynonymous | Exon 1 | c.424C>A | p.L142M |
| 3 | 121476641 | 121476641 | rs not annotated | G | C | exonic | *POLQ* | nonsynonymous | Exon 20 | c.6304C>G | p.Q2102E |
| 3 | 121923043 | 121923043 | rs753517272 | C | T | exonic;splicing | *SLC15A2* | stopgain | Exon 9 | c.778C>T | p.Q260X |
| 3 | 122568928 | 122568928 | rs150765058 | C | T | exonic | *DTX3L* | nonsynonymous | Exon 3 | c.839C>T | p.T280I |
| 3 | 123448538 | 123448538 | rs1279127415 | C | A | exonic | *ADCY5* | nonsynonymous | Exon 1 | c.8G>T | p.G3V |
| 3 | 123700313 | 123700313 | Rs1452968448 | T | C | exonic | *MYLK* | nonsynonymous | Exon 15 | c.3155A>G | p.N1052S |
| 3 | 125091448 | 125091448 | rs762284041 | G | A | exonic | *SLC12A8* | nonsynonymous | Exon 6 | c.1315C>T | p.L439F |
| 3 | 127003384 | 127003384 | rs199693063 | G | A | exonic | *PLXNA1* | nonsynonymous | Exon 3 | c.1432G>A | p.V478I |
| 3 | 128910310 | 128910310 | rs765706722 | G | A | exonic | *ACAD9* | nonsynonymous | Exon 8 | c.1192C>T | p.P398S |
| 3 | 128945350 | 128945350 | rs775891329 | C | T | exonic | *CFAP92* | nonsynonymous | Exon 3 | c.617G>A | p.R206H |

**Suppl. 3.5.** List of 17 coding variants (MAF < 0.01) under final investigation.

**Note:** **ZBTB20* variant present in all affected individuals in homozygosis
