## Supplementary figures and images for "Mutations in *ZBTB20* in individuals with persistent stuttering"

### Supplemental 3.6

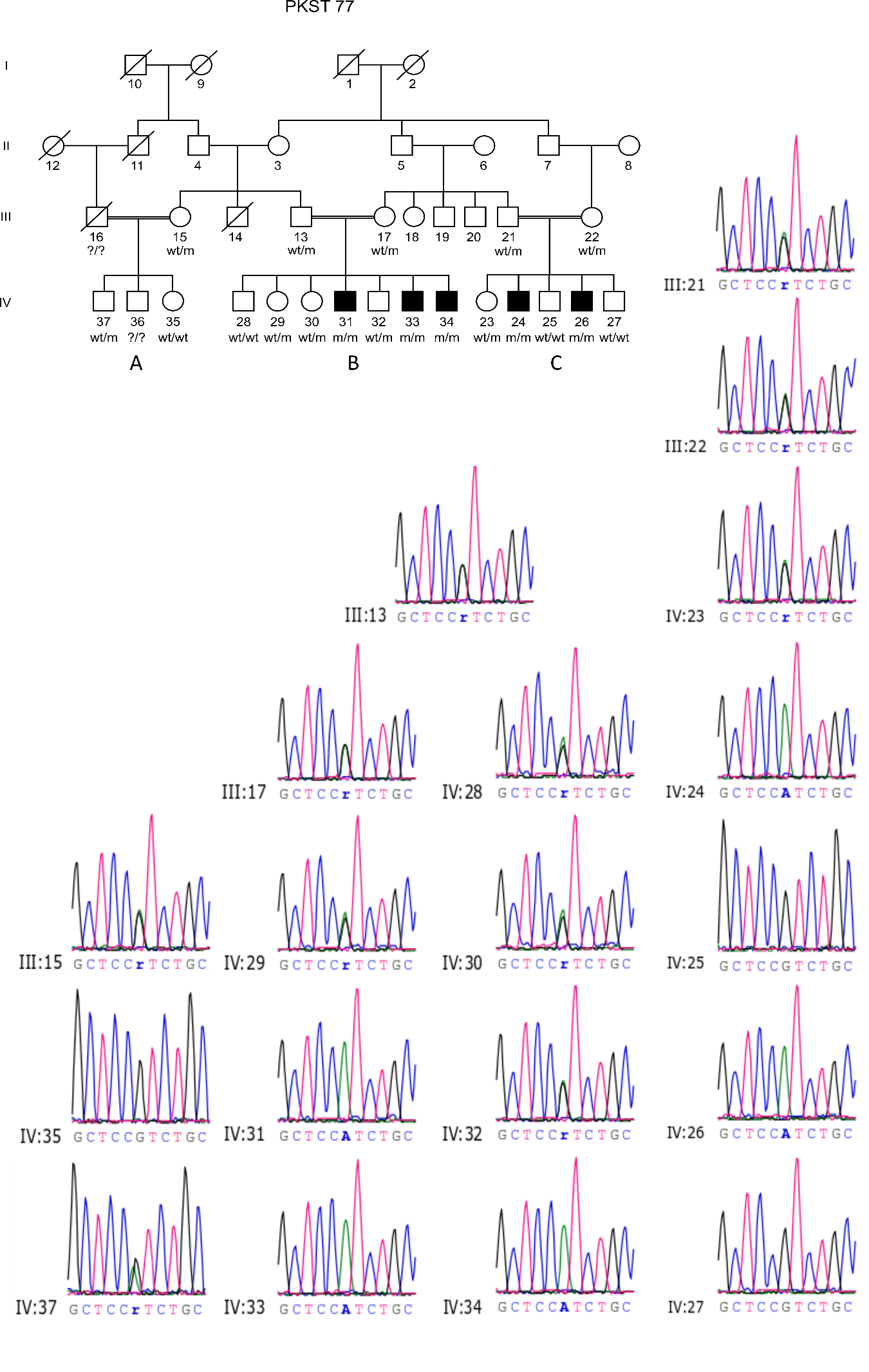
